# Disentangling the genetic and non-genetic origin of disease co-occurrences

**DOI:** 10.1101/2025.10.16.25338165

**Authors:** Beatriz Urda-García, Davide Cirillo, Alfonso Valencia

## Abstract

Numerous diseases co-occur more than expected by chance, likely due to a combination of genetic and environmental factors. However, the extent to which these influences shape disease relationships remain unclear. Here, we integrate large-scale RNA-seq data and heritability measures from human diseases with genomic data from the UK Biobank to disentangle the genetic and non-genetic origins of disease co-occurrences (DCs). Our findings show that gene expression not only recovers but also expands upon genomically explained DCs, capturing disease relationships beyond genetic variation. Approximately 60% of transcriptomically inferred DCs have a detectable genomic component, whereas the remaining 40% are not explained by known genomic layers, suggesting contributions from regulatory or environmental mechanisms. Consistent with this interpretation, the relative contributions of transcriptomics and genomics reconstruct disease etiology and correlate with comorbidity burden, revealing key aspects of disease mechanisms. Additionally, we find that diseases do not generally co-occur based on their heritability, except when sharing SNPs. However, highly heritable diseases tend to have genetically driven co-occurrences, even with lowly heritable diseases. In contrast, transcriptomics explains DCs regardless of heritability, at least partly due to non-heritable mechanisms, such as regulatory or environmental. Integrating transcriptomic and genomic data provides near-complete coverage of DCs among the analyzed diseases, with a considerable portion likely rooted in factors beyond DNA sequence and, therefore, potentially modifiable.

## Introduction

Strikingly, 1 out of every 3 adults suffers from two or more diseases worldwide, being this the norm for the elderly and even more pronounced for women [1] and socioeconomically deprived communities [2]. Contrary to intuition, there are actually more young than elderly people affected by this phenomenon worldwide, which considerably decreases life quality and expectancy, complicates the choice of therapy, and substantially increases health care cost [2–4]. Despite the massive prevalence and impact of disease co-occurrence, most health care is still designed to treat individual conditions.

In fact, numerous diseases tend to co-occur more than expected by chance. We will use the term disease co-occurrences (DCs) to refer to pairs of diseases that significantly co-occur in the same person either simultaneously (multimorbidities) or within a time window (comorbidities). Static and temporal epidemiological networks connecting all comorbid or multimorbid diseases of a given population have been built [5–7], revealing some subgroup-specific patterns [6].

The observed patterns suggest that DCs, like diseases themselves, may result from the convergence of genetic and environmental factors that ultimately lead to alterations in specific molecular mechanisms. These alterations can be inherited or acquired, and may involve genetic variants, epigenetic modifications, transcriptional, or post-transcriptional changes, as well as their interplay with environmental factors. Understanding the relative contributions of these genetic and environmental influences on diseases and their co-occurrences remains a fundamental question in biology, which this manuscript aims to address.

Deciphering why diseases co-occur is crucial for improving prevention, diagnosis, and treatment of patients. To this end, a myriad of studies focused on finding the molecular basis of specific DCs [8–10]. Subsequently, systematic studies successfully showed that DCs are enriched in a variety of molecular components (e.g., they tend to share disease-associated genes [11, 12], pathways [12], coupled metabolic reactions [13], and are closer in PPI networks [14]). Yet, systematic methods recovered a small percentage of the epidemiology among their analyzed diseases (*<*9% except microarrays [15] with 16%) [16]. Recent advances suggest that transcriptomic data may provide a more comprenhensive explanation for disease co-occurrences, raising questions on interpretability, as the transcriptome can potentially capture a diverse array of factors, ranging from genetic to environmental influences [16]. In parallel, Dong *et al.*, used genetic and epidemiological data from the UK Biobank (UKB) to derive a multimorbidity network, providing the disease pairs that share a genomic cue, such as SNPs and other molecular components hierarchically inferred from them (disease-associated genes and pathways, PPIs containing disease-associated genes and genetic correlation) [17, 18].

In this study, we integrated RNA-seq data on human diseases and genomic data from the UKB to disentangle the genetic and non-genetic origin of disease co-occurrences. First, we built largescale disease and stratified networks based on the transcriptomic similarities of diseases and their molecular subgroups, respectively. By systematically analyzing transcriptomic and genomic layers, we examined their contributions to disease co-occurrences and explored the molecular dependencies shaping these interactions. As part of this, we quantified the proportion of transcriptomically explained disease co-occurrences with a genomic basis versus those potentially driven by regulatory or environmental influences on expression. We further examined the molecular landscape of disease co-occurrences and its relationship with comorbidity burden. Finally, we leveraged recent large-scale heritability measures for human diseases [19] to assess the relationship between disease heritability and the molecular underpinnings of their co-occurrences, offering new insights into their genetic and environmental influences.

## Results

### Transcriptomic and genomic contributions to DCs

In this study, we aim to determine the proportion of transcriptomic similarities observed in disease co-occurrences that can be attributed to genetic factors versus those potentially influenced by environmental effects. This approach enables us to explore the relationship between genomic factors identified from UKB data and gene expression patterns derived from RNA-seq, which serve as a proxy for the interplay between genetics and the environment. As expected, we observed overlaps between genomic components and gene expression, alongside partial differences reflecting the additional impact of environmental or beyond genetic influences on expression.

First, we leveraged large-scale RNA-seq data sets to build two networks based on the transcriptomic similarities and dissimilarities of human diseases: a Disease Similarity Network (DSN) linking diseases, and a Stratified Similarity Network (SSN) that connects both diseases and subgroups of patients with similar gene expression profiles (Methods). Both the DSN and the SSN consist of a single connected component, with a predominance of positive interactions (∼60%)(Note S1, Table S1). These networks significantly recall a robust and considerable proportion of known disease co-occurrences in the gold standard [5] and the independently generated UKB epidemiological network [18] (Methods, Table S2, Note S2).

The UKB provides the shared genomic components for the DCs in the UKB epidemiological network (UKB DCs) (Methods, Note S3). Therefore, we compared the overlap of the disease and stratified RNA-seq networks (DSN and SSN) with the genomic layers over the UKB DCs for the common set of diseases (97 ICD10 codes). We show the results before and after filtering for the diseases containing GWAS summary statistics in the UKB (Methods).

48.25% of UKB DCs were captured by the DSN, followed by pathways (29.7%), genetic correlation (24.28%), genes (22.36%), PPIs (22.04%), and SNPs, with a small 3.35% (Fig. 1a). Interestingly, the joint recall of the genomic layers is 45.85%, similar to the recall of the transcriptomic layer alone. When inspecting how the DCs distribute between the different layers, we clearly see that the most frequent scenarios by far are unexplained DCs (DCs not explained by any molecular layer) and DCs explained solely by the DSN (Fig. 1c). Analogous patterns are observed when filtering diseases without GWAS (Fig. 1b-d). In both cases, the third position comprises DCs explained by both the DSN and genetic correlation (Fig. 1c-d). Moreover, only 11 interactions can be explained by all 4 UKB layers, 7 (63.6%) of which are also captured by the DSN.

**Figure 1:**
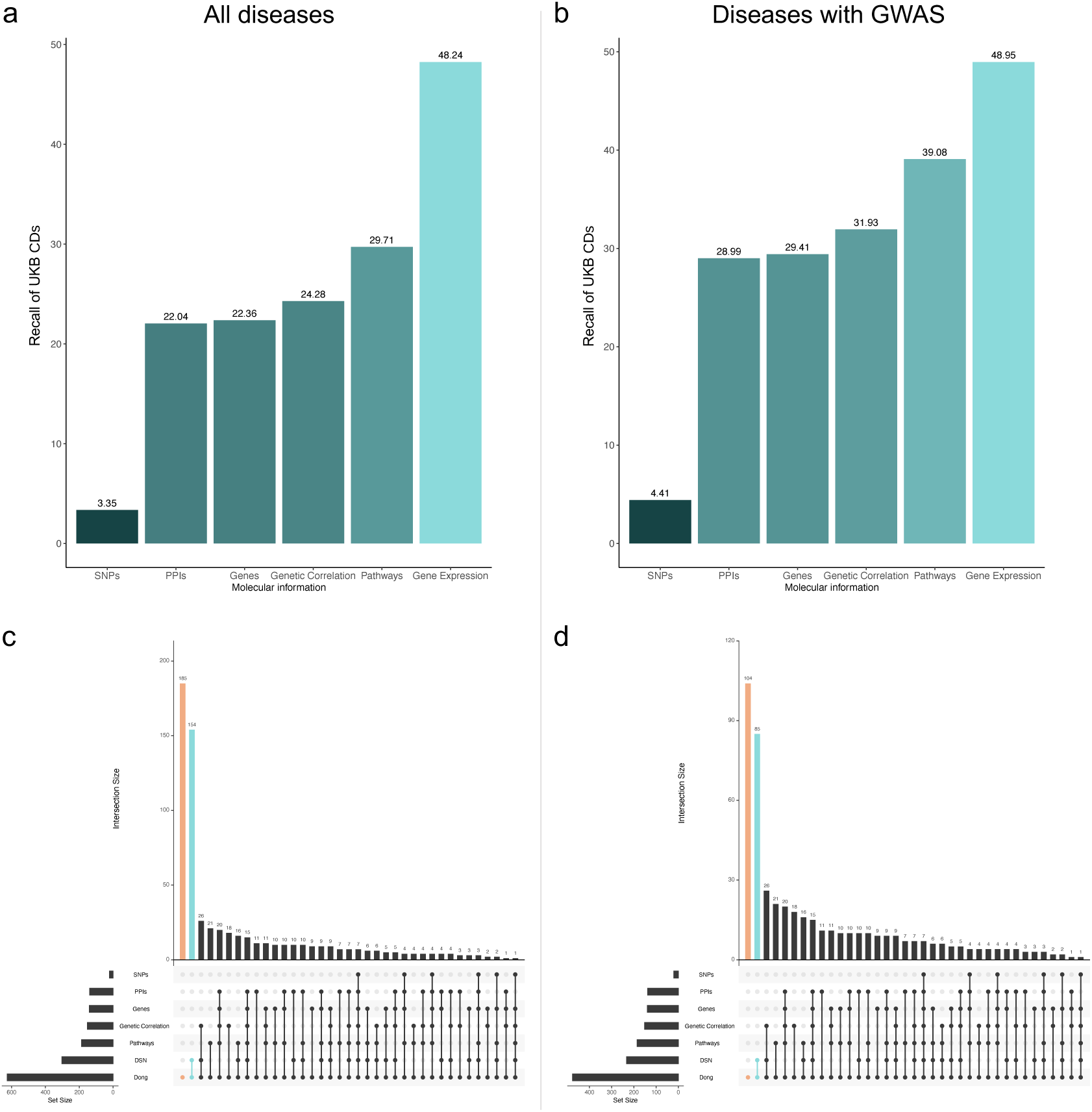
Comparison between the recalls of the Disease Similarity Network (DSN) and the genomically-derived layers. (a-b) Recall of UKB disease co-occurrences (DCs) for the different molecular layers for the common set of diseases (a) and for the subset of diseases containing GWAS summary statistics (b). (c-d) Upset plots of the distribution of the UKB DCs among the molecular layers for the common set of diseases (c) and the subset containing GWAS (d). In it, unexplained DCs and DCs captured solely by gene expression are colored in orange and light green respectively.

Similar yet more extreme behavior is observed for the stratified network (SSN), which far exceeds the ability of the genomic layers to capture DCs, significantly recalling a robust 77.48% and 77.73% of DCs before and after the disease filtering (Fig. 2a-b). DCs explained solely by the SSN now constitute the single most frequent scenario, followed by unexplained DCs and DCs explained by both SSN and genetic correlation, both before and after disease filtering (Fig. 2c-d).

**Figure 2:**
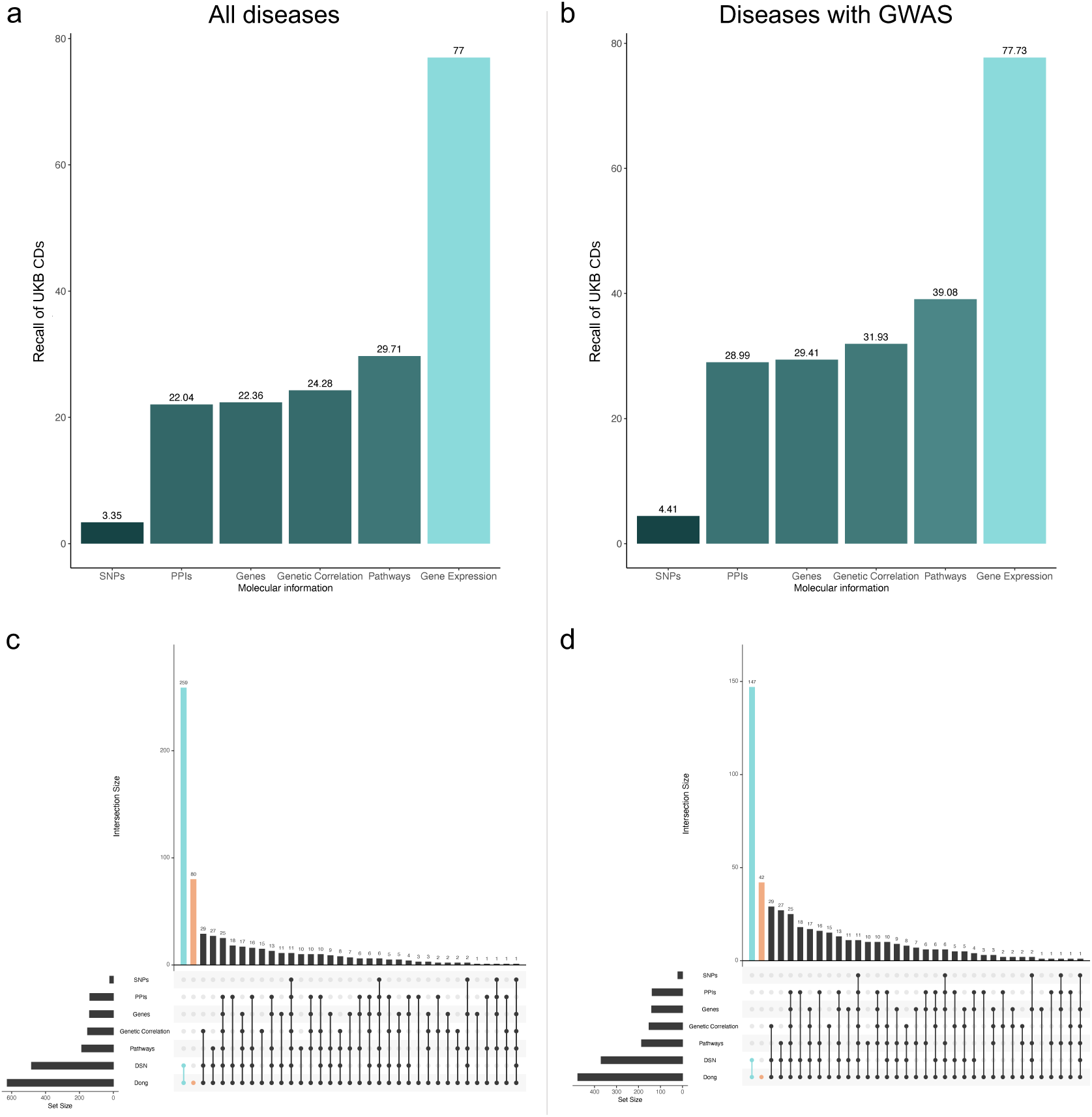
Comparison between the recalls of the Stratified Similarity Network (SSN) and the genomically-derived layers. (a-b) Recall of UKB disease co-occurrences (DCs) for the different molecular layers for the common set of diseases (a) and for the subset of diseases containing GWAS summary statistics (b). (c-d) Upset plots of the distribution of the UKB DCs among the molecular layers for the common set of diseases (c) and the subset containing GWAS (d). In it, unexplained DCs and DCs captured solely by gene expression are colored in orange and light green respectively.

In all, a total of 87.2% of DCs can be explained by molecular information, where 45.85% is captured by at least one of the genomic layers and 41.4% is uniquely captured by gene expression. Furthermore, 93.3% of DCs are captured if we consider diseases with GWAS, leaving unexplained less than 7% of comorbidities for which genomic and transcriptomic information is available. This shows that combining genomic and transcriptomic layers provides almost complete coverage of the DCs among the covered diseases, being gene expression the single most informative layer (*>*77% recall).

### Identifying genomic signatures in RNA-seq associations across DCs

The biological relationships between the different molecular components and the way in which the UKB genomic layers have been derived (hierarchically from SNPs) (Fig. 1a) suggest that there might be dependencies between the molecular layers and lead to the following question: where does RNA-seq fit?

To explore this, we have used a directed measure of mutual information I(X,Y)/H(X) that provides the proportion of information of X that can be explained by Y, where each variable encodes the DCs in the UKB captured by a given molecular layer (Fig. 3c, Methods). The analysis reveals that gene expression is independent of the rest of the variables, both at the disease and stratified level (Fig. 3a-b). The remaining layers vary on their dependence level, not necessarily in a symmetric manner. For instance, even though genes are derived from SNPs, 31% of the information derived from SNPs can be explained by genes (compared to the 9% in the opposite direction) and, as expected, these numbers decrease with the subsequent layers. Smaller and more symmetric dependencies around 20%, 15%, and 14% are observed for genes–pathways, PPIs–pathways and genes–PPIs, respectively. Genetic correlation emerges as the most independent genomic layer, explained at most 10% by genes.

**Figure 3:**
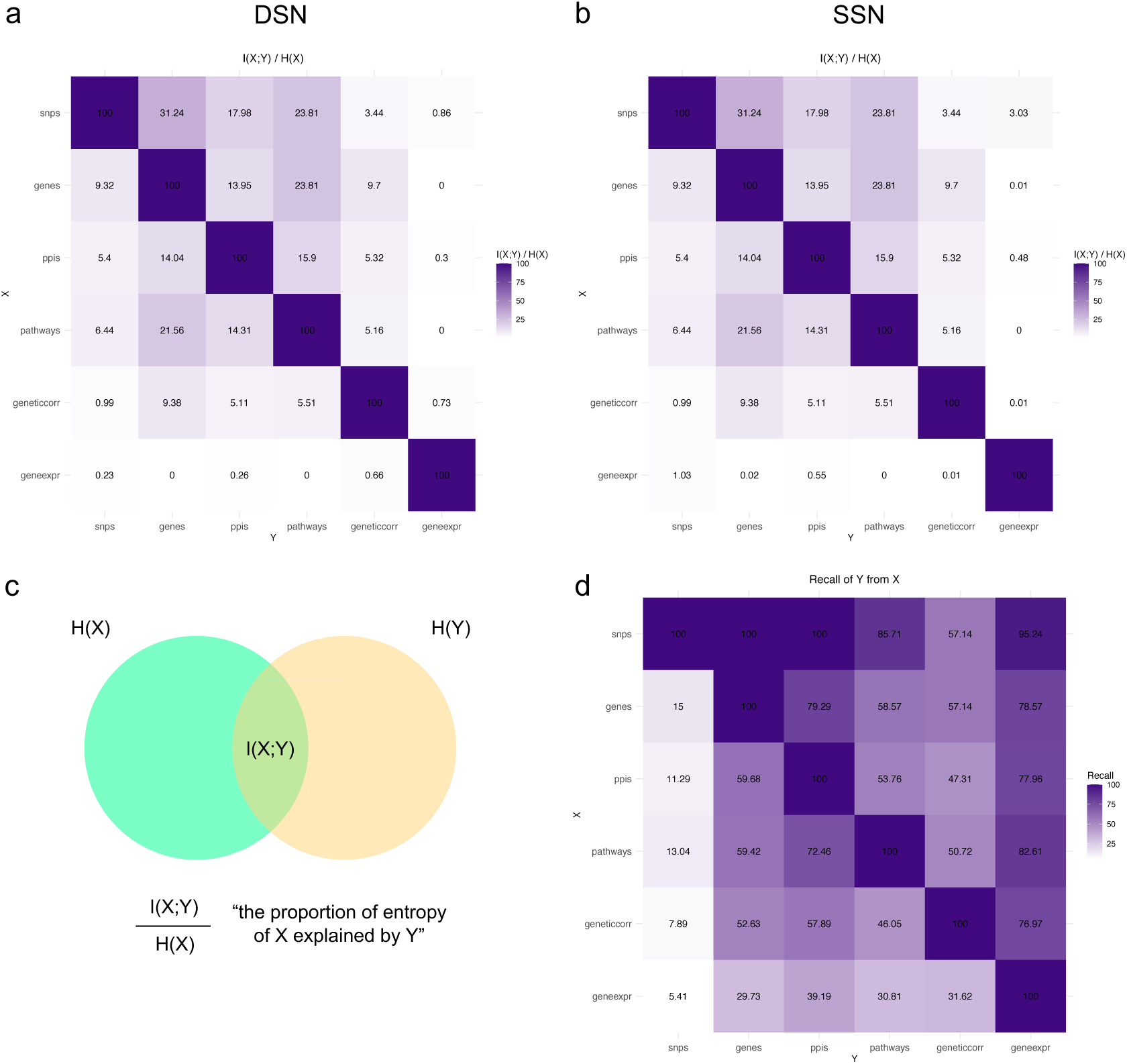
Dependency between the molecular layers. (a-b) Heatmap showing the proportion of mutual information of each molecular layer explained by the rest of them, considering for gene expression (a) the Disease Similarity Network (DSN) and (b) the Stratified Similarity Network (SSN) respectively (Methods). Values range from 0 (independence) to 100 (dependence). (c) Schema of the metric used to measure variable independence: I(X;Y)/H(X), where H(X) is the entropy of variable X and I(X;Y) is the mutual information of the variables X and Y. (d) Pairwise recall of all molecular layers over the common set of diseases. It shows the recall of variable Y (columns) of the disease co-occurrences explained by variable X (rows).

When considering the Jaccard index, the gradient from SNPs to genes, pathways, PPIs, genetic correlation, and gene expression remains (Fig. S1). High levels of overlap are observed between genes, PPIs, and pathways (e.g. 52% of the DCs captured by either genes or PPIs are contained by both), followed by genetic correlation and gene expression (means of 28% and 22%). Actually, gene expression recalls from 77 to 95% of the interactions explained by the genomic layers, and adds many more missed by them, broadening the scope of genomic data (Fig. 3d).

These results show that gene expression covers the vast majority of genomically interpretable DCs, establishing that a significant portion (60%) of its DCs have a genetic basis. Moreover, gene expression captures many additional disease co-occurrences, accounting for its observed independence and reflecting its ability to integrate influences beyond genetics.

### Molecular landscape of disease co-occurrences

After confirming that nearly all disease co-occurrences have a molecular queue, we can explore how diseases distribute in this molecular space. We mapped diseases based on the relative efficacy of transcriptomics versus genomics in capturing their co-occurrences (Fig. 4). We observe that most diseases fall into the first two quadrants, indicating that diseases tend to be reasonably well captured by transcriptomics while exhibiting a wide range of genomic contributions. Next, we will explore different scenarios within this landscape.

**Figure 4:**
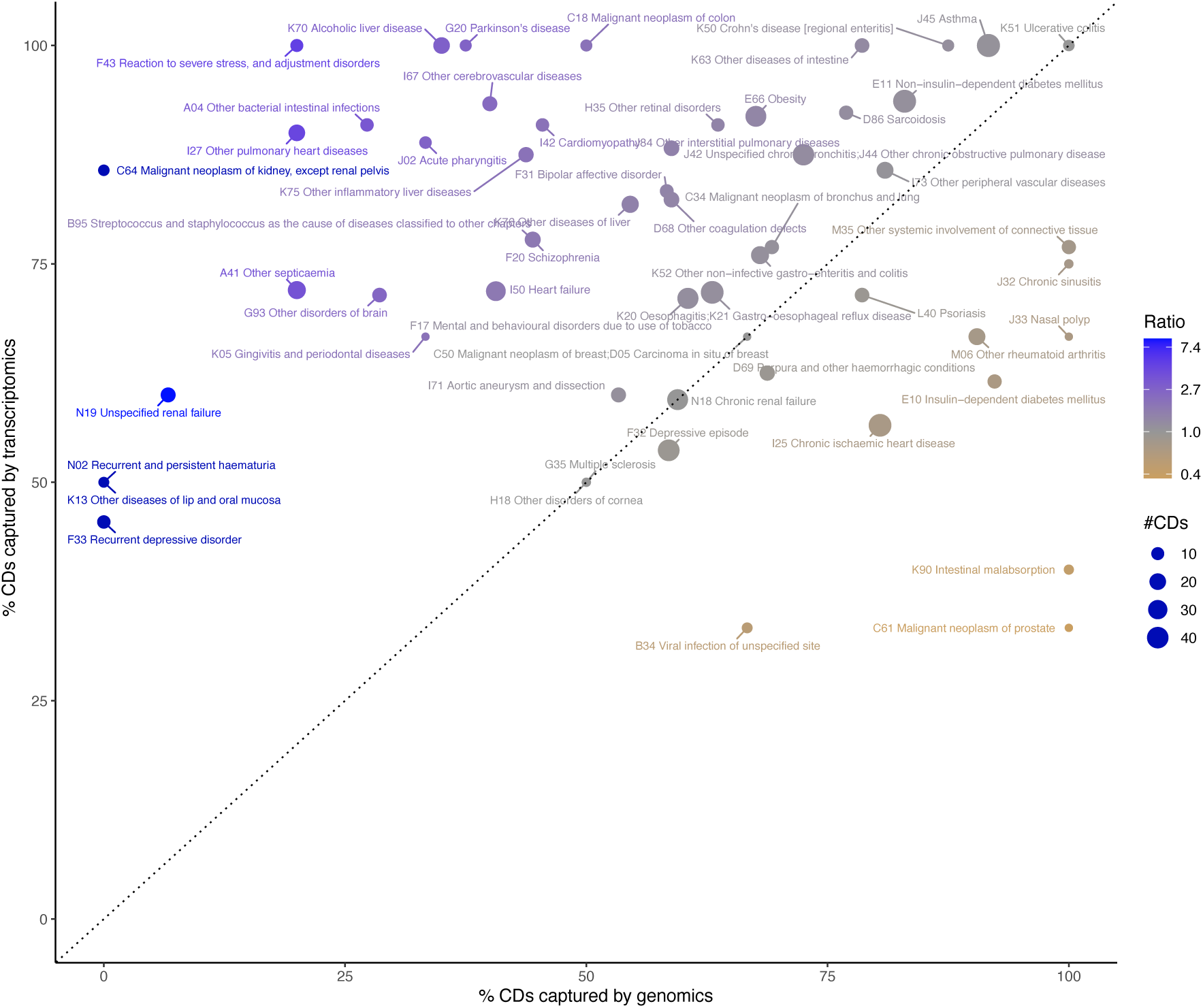
Transcriptomic and genomic contributions to disease co-occurrences (DCs) of human diseases. Scatter Plot illustrating the transcriptomic and genomic contributions to DCs for each disease. Each point represents a disease, with the percentage of DCs explained by transcriptomics on the y-axis and genomics on the x-axis. Point size corresponds to the number of DCs associated with each disease in the UK Biobank epidemiological network [18]. Diseases better explained by genomics are depicted in yellow, while those better explained by transcriptomics are shown in blue. Dark blue points represent diseases whose DCs are exclusively explained by transcriptomics.

Various diseases are well-characterized by both transcriptomic and genomic data, underscoring the complementary roles of these molecular approaches in understanding disease co-occurrences. One of these diseases is ulcerative colitis (UC), the most common form of inflammatory bowel disease worldwide. A family history of UC is the strongest risk factor for the disease, with first-degree relatives of patients having a fourfold higher risk of developing the disease [20]. Besides genetic predisposition, environmental factors heavily influence UC. For instance, a westernized lifestyle is a known risk factor for both the incidence and severity of UC, whereas smoking has a protective effect against both the development and severity of it [20–23]. We observe that both omics effectively capture UC disease co-occurrences (Fig. 4). The co-occurrence of UC with Crohn’s disease and other gastrointestinal disorders (e.g., gastroesophageal reflux disease) points to common inflammatory processes influenced by shared genetic susceptibilities and immune responses. The co-occurrence with chronic ischemic heart disease, asthma and type 2 diabetes underscores the systemic impact of chronic inflammation. Indeed, UC shares 5 to over 50 susceptibility genes with these conditions, many of which are located in the Major Histocompatibility Complex (e.g. *HLA-B* ;*HLA-C*) or are related to the immune system. Key genes underlying these interactions also include gene expression regulators via histone methylation (*EHMT2*) or RNA processing (*DDX39B*). Notably, we were also able to detect the inverse relationship between UC and smoking at the transcriptomic level. It is well-established that UC is more prevalent among nonsmokers or individuals who recently quit smoking. Additionally, smokers diagnosed with UC tend to exhibit milder disease, requiring fewer hospitalizations and reduced medication [20].

Asthma is one of the most common chronic conditions, with a widely accepted strong hereditary component from 55–74% in adults, and almost reaching 90% in children [24, 25]. Indeed, 44 out of the 48 co-occurrences of asthma with other diseases have a genomic commonality, mostly influenced by sharing risk genes, derived network components or significant genetic correlations (Fig. 4, 5e). Even so, asthma is highly heterogeneous and results from a complex interplay of gene-environment interactions, where genetic susceptibility, epigenetic and environmental factors determine the actual development and progression of the condition [26]. Air pollutants, allergens, intake of several drugs and suplements, obesity and even maternal and paternal diseases and lifestyle have been linked to its incidence [27, 28]. Strikingly, asthma incidence has rapidly increased by 3-5 fold in 40 years while being highly heritable, pointing to the existence of heritable factors beyond DNA sequence [24]. We observe that all 48 DCs of asthma are significantly captured by transcriptomics, 54% of which were only revealed at the stratified level, unveiling the hetereogeity of the disease also regarding comorbidities.

Other diseases whose comorbidities are well captured by both omics and that are known to result from an interplay of strong genetic factors and environmental exposures include sarcoidosis [29–31] and rheumatoid arthritis (Fig. 4) [32, 33]. On the other hand, intestinal malabsorption, often rooted in immune and autoimmune genetic susceptibilities or congenital defects that tend to be monogenic, exhibits fewer links that are generally better explained by genomics (Fig. 5f) [34].

**Figure 5:**
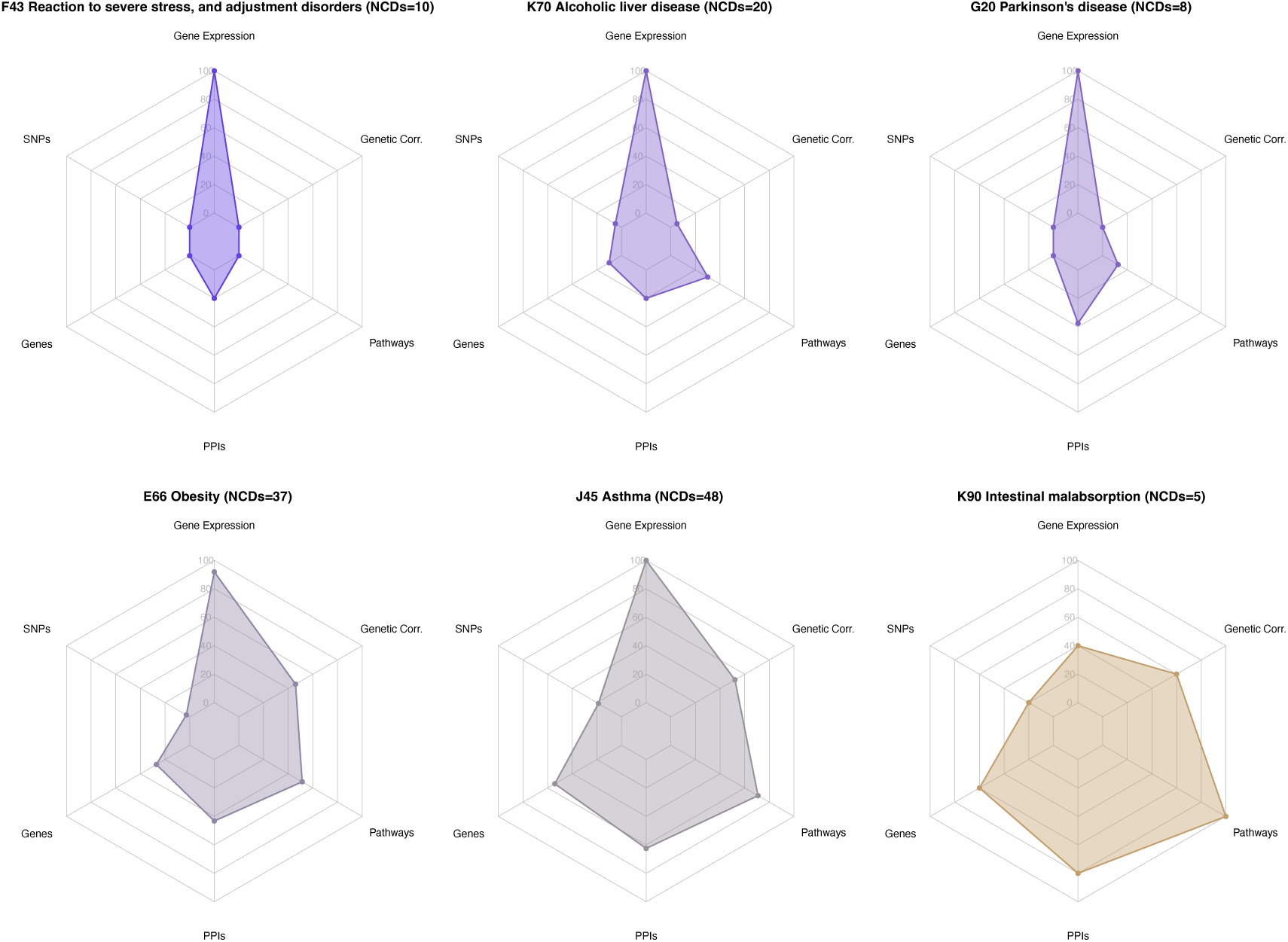
Radar charts illustrating the molecular contributions to disease co-occurrences (DCs) of diseases. Each radar chart presents the ability of six distinct molecular layers–gene expression, genetic correlation, pathways, protein-protein interactions (PPIs), genes, and single nucleotide polymorphisms (SNPs)–in capturing the DCs associated with a specific disease. The axes of the radar charts represent the recall of each molecular layer, indicating its ability to explain DCs. Longer axis lengths signify a higher recall, suggesting a greater influence of the corresponding molecular layer on the DCs observed for the disease. Diseases are colored based on the ratio of the transcriptomic versus overall genomic recall, with diseases better explained by transcriptomics depicted in blue, those better explained by genomics are shown in yellow and the ones with equal transcriptomic and genomic recall appear in grey. ICD10 code, disease name, and number of interactions in UKB epidemiological network (NCDs) are disclosed.

Diabetes serves as an illustrative example. Type 1 diabetes (T1D, ICD10=E10) is caused by the autoimmune destruction of pancreatic beta cells, leading to reduced insulin production. Conversely, type 2 diabetes (T2D, ICD10=E11) is characterized by insulin insensitivity, primarily caused by insulin resistance or inadequate insulin production [35]. While both diseases are in the second quadrant, T1D is more effectively captured by genomics, whereas T2D is better represented by transcriptomics (Fig. 4). Indeed, T1D typically manifests at a younger age and has a stronger genetic component, with heritabilities around 56% for adults and up to 81% for children [36], largely attributed to susceptible loci, particularly in the HLA region [37]. Nevertheless, epigenetic and environmental triggers, such as cold weather, air pollution, certain viruses, or early diet, have been identified and recently associated to the rising incidence of the disease [37, 38]. On the other hand, T2D usually appears later in life and, despite having a strong genetic component, it is more complex and poorly explained by the many identified variants [35, 37]. In this case, environmental factors including diet, exercise, stress, and sleeping patterns are main determinants of the disease [35].

Other diseases are well captured by transcriptomics and moderately by genomics. This is the case for obesity, which actually results from an intricate interplay of genetic predispositions and strong environmental influences. 92% of obesity co-occurrences with other diseases are significantly captured by transcriptomics and 68% share a genomic queue, with a low contribution of SNPs and genes (Fig. 4, 5d). Both omics capture associations with T2D, cardiomyopathy, heart failure, aortic aneurism, asthma, chronic bronchitis or breast cancer. Here, genetic correlation and pathways are the main genomic queue, aligned with the complex nature of the disease itself. Many co-occurrences are only captured by gene expression, including colorectal and kidney cancer, renal failure, schizophrenia, recurrent depressive disorder, or reaction to severe stress or adjustment disorders, among others. As an example, colorectal cancer is widely regarded as an environmental disease involving a wide range of cultural, social and lifestyle factors. Most cases of colorectal cancer occur in people with no family history of it, with risk factors including an unhealthy diet high in animal fat and red meat and low in fiber, smoking, physical inactivity and even low socioeconomic status [39, 40].

Finally, numerous diseases are better characterized by transcriptomics, with modest to none genomic queues underlying their disease co-occurrences (Fig. 4). Diseases in this area are mostly due to or considerably affected by factors beyond DNA sequence. Examples include reaction to severe stress, and adjustment disorders (PTSD) (five times better explained by transcriptomics, Fig. 5a), alcoholic liver disease (Fig. 5b), mental and behavioral disorders due to use of tobacco (smoking), or the previously mentioned colorectal cancer. We also see oral dysplasia (K13), commonly caused by smoking or alcohol consumption [41]; gingivitis and periodontal disease, generally linked to poor oral hygiene, smoking, nutritional deficits, or certain drugs [42]; or kidney cancer, with major risk factors including smoking, chemical exposure, or obesity, and with only 3-5% of cases occurring within a familial context [43].

Interestingly, most neoplasms are better explained by transcriptomics, with few exceptions such as breast cancer, widely known for its genetic contribution and equally captured by both omics (Fig. 4, S4) [44]. This is aligned with recent evidence showing that 60%–90% of all cancer cases are due to exogenous (e.g. lifestyle) and cell extrinsic risk factors (e.g. oxidative stress, microbiome, immune responses) that therefore could be modifiable [45]. Indeed, up to 30, 35 and 20% of cancer cases are linked to tobacco, diet, and infections, respectively [46]. Large population studies have shown that at least 40% of cancer cases are attributed to different combinations of these modifiable risk factors, including smoking, alcohol consumption, stress, physical activity, air pollution, or class I carcinogens [47–51]. In fact, oncogenic alterations can be inherited or acquired but they must operate in a permissive environment to drive cancer formation. Risk factors can also lead to the development of driver mutations, with specific tobacco-associated signatures in lung cancer or UV radiation signatures in melanoma [45].

Infectious diseases (e.g. bacterial infections, streptococcal and staphylococcal infections, septicaemia, or acute pharyngitis) are mostly explained by transcriptomics, with no SNPs and less than average genes, pathways and genetic correlations underlying their co-occurrences with other diseases (Fig. 4, S4). Indeed, recent studies show that SNP heritability for infection susceptibility is around 7% [52].

We also observe this pattern for most psychiatric and neurological diseases, including PTSD, parkinson’s disease, bipolar disorder, schizophrenia, smoking, cerebrovascular diseases, other diseases of brain or recurrent depressive disorder (Fig. 4, S4). Even though both omics underlie some of their connections, most are uniquely revealed at the level of gene expression. Only transcriptomics recalls (1) parkinson’s association with heart failure, oesophagitis, chronic bronchitis, or depression (Fig. 5c); (2) bipolar disorder co-occurrence with smoking, depression, PTSD, oesophagitis, or chronic renal failure; (3) recurrent depressive episode association with asthma, obesity, T2, or PTSD; or (4) smoking’s link to coagulation defects, kidney cancer, inflammatory liver diseases, gingivitis and periodontal disease, pulmonary heart diseases, depression, PTSD, schizophrenia, or multiple sclerosis.

To sum up, we mapped diseases based on the ability of transcriptomics and genomics to recall their epidemiological interactions with other diseases. We observed that diseases effectively captured by both omics generally have significant genomic contributions whereas those whose etiology is more affected by factors beyond DNA sequence tend be better captured by gene expression.

Expanding on this, we observed considerable variation in the number of co-occurring conditions across diseases (Fig. 4). We detected a small significant relationship between the number of co-occurring diseases and the underlying molecular burden, with highly comorbid diseases exhibiting more balanced contributions from both genomics and transcriptomics in their interactions (Kendall’s tau, p-value=0.013; quadratic regression, p-value = 0.024) (Methods, Fig. S5). In contrast, diseases with co-occurrences primarily supported by one of the layers tend to have fewer comorbidities. For instance, asthma, influenced by strong genetic and environmental contributors, has extensive comorbid conditions (48 co-occurrences) supported by molecular data in a balanced manner (Fig. 4, 5). In comparison, often monogenic conditions such as intestinal malabsorption, which may exert more localized effects without broad changes in gene expression patterns, show fewer co-occurrences (5 co-occurrences) primarily supported by genomics. Actually, genetic correlation is the only genomic layer whose recall shows significant association with the number of comorbidities (Kendall’s tau=0.2, p-value=0.0186) (Methods). This finding suggests that cumulative, small genetic effects across the genome may have a greater impact on multidisease burden than isolated mutations affecting specific genes or biological processes. Together, these results suggest that a diverse and multilayered molecular basis may enhance the likelihood of comorbidities, likely due to broader disruptions in both local and systemic mechanisms.

### Highly heritable diseases tend to share genomic components with their co-occurring diseases

The genetic component of diseases can be disentagled by measuring their heritability (*h*^2^)–the proportion of phenotypic variance attributable to inherited genetic factors. Here, we used recent heritability measures on human diseases based on nationwide Danish pedigrees to evaluate the relationship between disease heritability and the molecular bases of their co-occurrences with other diseases (Methods) [19].

First, we computed the assortativity based on *h*^2^ to evaluate if diseases have a tendency to co-occur with diseases with similar heritability values (Methods, Fig. S7). We did not observe this tendency for overall DCs or DCs captured by any of the molecular layers except for SNPs. Therefore, DCs explained by SNPs tend to involve diseases with similar heritabilities. Nonetheless, DCs and DCs explained by gene expression or other genomic components are not more frequent between diseases with similar heritable components.

Secondly, we observe that disease heritability significantly correlates with the proportion of co-occurring diseases explained by genomics (Methods, Fig. 6). In other words, highly heritable diseases tend to have disease co-occurrences with a genomic component, whereas fewer genomic components are found in the co-occurrences of lowly heritable conditions. This is true for co-occurrences sharing SNPs, PPIs, pathways, significant genetic correlations, and any genomic layer (genomic) (Fig. 6).

**Figure 6:**
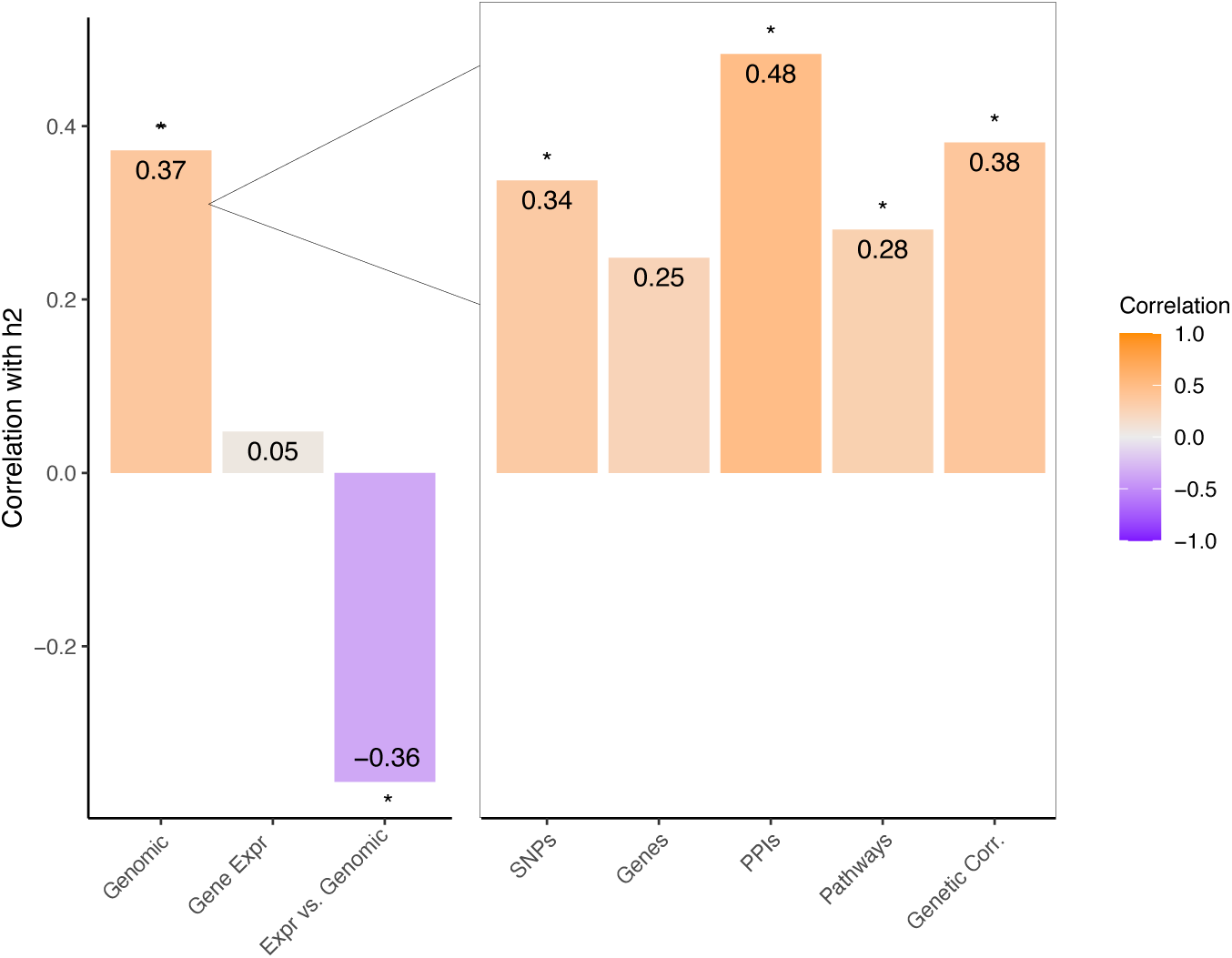
Correlation of Heritability with the Proportion of Disease Co-occurrences Captured by Molecular Layers. This figure illustrates Spearman’s correlation between heritability (h2) and the proportion of disease co-occurrences (DCs) captured by genomics (any genomic layer), gene expression, and the ratio between gene expression and genomics (Expr vs. Genomics) (Methods). It also shows the correlation for each genetic layer: SNPs, genes, protein-protein interactions (PPIs), pathways, and genetic correlation. Significant correlations are marked with an asterisk. The results show that all genomic layers, except for genes, have a significant correlation between the heritability of the diseases and the ability of genomics to capture their co-occurrence with other diseases. Gene expression’s recall does not correlate with heritability, and the ratio between gene expression and genomic layers shows a significant negative correlation.

These results indicate that (1) generally, genetic diseases do not tend to associate with other genetic diseases and (2) genetic diseases tend to share genomic components with their co-occurring conditions, even if these are not highly heritable. For instance, asthma, a highly heritable condition, tends to co-occur with highly heritable diseases such as ulcerative colitis (supported by most genomic components) and with lowly heritable conditions such as several infections (with some genes underlying the interactions).

On the other hand, disease heritability does not correlate with the ability of transcriptomics to capture their co-occurrences (Fig. 6). This aligns with the observation that transcriptomics capture the disease co-occurrences of both highly and lowly heritable diseases in a similar manner. Interestingly, the ratio between the transcriptomic and genomic recall is negatively correlated with heritability, indicating that at least a proportion of the recall solely captured by transcriptomics accounts for non-heritable factors, such as environmental influences, gene environment interactions or other disease-associated molecular mechanisms, and not only heritable factors beyond DNA sequence.

Spousal heritability 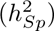–the proportion of variance attributable to shared spousal environmental components–serves as a proxy for the environmental component of diseases (Methods) [19]. Table S5 shows the top 15 diseases with the highest spousal heritability, which, as previously discussed, are mostly explained by transcriptomics with moderate to none genetic queues. The top diseases are PTSD, smoking, and obesity, followed by various infectious diseases (e.g., streptococcal and staphylococcal infections and other bacterial infections), psychiatric diseases (e.g., schizophrenia), and other diseases with strong environmental influence and transcriptomic recall, such as alcoholic liver disease and T2D (Fig. S8).

## Discussion

Diseases generally arise from a convergence of genetic and environmental influences that ultimately lead to molecular disruptions. Some conditions result from direct genetic mutations, as seen in rare diseases, while others emerge from complex interactions between genetic susceptibilities and external factors. Understanding the relative contributions of these influences is fundamental to disease biology, yet remains a major challenge. The same complexity applies to disease co-occurrences, which offer a unique opportunity to explore this balance. In this context, genomic and RNA-seq data serve as key tools for disentangling the genetic and non-genetic drivers of disease relationships.

Despite inherent limitations, transcriptomic data captures a significant portion of disease co-occurrences, raising an important question: do these similarities primarily reflect a shared genetic basis, or are they driven by other factors, such as environmental influences? To investigate this, we leveraged genomic data from the UK Biobank (UKB) analyzed by Dong *et al.* and found that approximately 60% of disease co-occurrences identified through transcriptomics also exhibit a detectable genomic component, such as shared SNPs, genes, genetic correlations, or other molecular network elements. Furthermore, gene expression captures the majority of genetically driven DCs, covering up to 95% of those with a known genomic basis, reinforcing its value as a proxy.

However, the remaining 40% of the gene expression DCs were missed by the genomic layers, suggesting that many DCs might share molecular mechanisms not rooted in mutations affecting the same gene, interacting proteins, or pathways. Indeed, our findings indicate that gene expression operates largely independently of other genomic layers, capturing unique aspects of disease biology not fully explained by the available genomic data. This could include the effect of known and unknown genetic variants, epigenetic, transcriptional, or post-transcriptional changes, and both the effect and interplay of these mechanisms with environmental factors.

Consistent with this interpretation, mapping diseases based on the genomic and transcriptomic contributions to their co-occurrences reconstructs disease etiology, uncovering key aspects of disease mechanisms. Complex diseases with strong genetic contributions, such as ulcerative colitis and asthma, are robustly captured by both omics, underscoring their complementary roles. Diseases characterized by a complex interplay of genetic susceptibilities and environmental or epigenetic factors align well with their known etiologies in our mapping. For example, T1D, with its strong genetic basis, is more accurately recalled by genomics, whereas T2D, which is more influenced by lifestyle and environmental factors, is better captured by transcriptomics. Obesity, a condition with primary environmental contributions coupled with genetic predisposition, exhibits strong recall by transcriptomics and moderate recall by genomics, highlighting the importance of both factors in its development. Cancers, particularly those with substantial environmental triggers, are predominantly explained by transcriptomics. Moreover, diseases primarily driven by environmental factors—such as PTSD, alcoholic liver diseases, infections, smoking, or numerous cancers—tend to exhibit high transcriptomic recall with little to no genomic contributions, underscoring the significant influence of non-genetic factors. This mapping approach not only reveals the unique etiologies underlying the co-occurrences of individual diseases but also provides valuable insights into the broader understanding of disease mechanisms.

Building on these findings, we observed that highly comorbid diseases tend to exhibit similar molecular contributions from genomics and transcriptomics, whereas diseases with fewer co-occurrences are often predominantly associated with one of these layers (Fig. S5). These observations suggest that diverse and multilayered molecular disruptions, including small cumulative genetic effects across the genome and transcriptomic alterations, may enhance the likelihood of disease co-occurrences.

The exploration of heritability further clarifies the relationship between genetic predispositions and disease co-occurrences. Our findings indicate that diseases do not tend to co-occur with others that have similar heritability levels, with a notable exception: co-occurrences sharing SNPs (e.g. ulcerative colitis with Crohn’s disease, diabetes or asthma). This suggests that, as expected, SNPs may play a unique role in linking diseases with similar genetic predispositions. Additionally, we observed a significant correlation between disease heritability and the genomic recall of disease co-occurrences. This indicates that highly heritable diseases do not tend to associate with highly heritable diseases; instead, they are more likely to have co-occurrences that are genetically driven, while diseases with lower heritability exhibit fewer genetic contributions in their co-occurrences. Conversely, transcriptomics captures co-occurrences across both highly and lowly heritable diseases with comparable efficacy, where at least a portion of its recall could be attributed to shared nonheritable molecular mechanisms.

These findings underscore the complementary roles of genomic and transcriptomic data in unraveling the complex interplay between heritability, environmental influences, disease etiology, and disease co-occurrences. It is estimated that 70-90% of disease risk is attributable to environmental exposures [53, 54]. These exposures, along with genetic predispositions, epigenetic modifications, and microbiome compositions, collectively shape the transcriptome, ultimately leading to the development of multiple, often interconnected diseases. Although many diseases are considerably heritable, known variants typically explain less than 10% of their risk [55, 56], suggesting the effect of unknown rare or structural variants, or environmental modulation on genetic effects though gene–environment interactions, epigenetic modifications, gene–gene interactions or shared exposure to environmental queues. Our findings suggest that the transcriptome may bridge this gap, capturing interactions that genomic data alone cannot fully explain (Fig. 4, Fig. S9). Indeed, these mechanisms have been shown to underlie both diseases and their co-occurrences [57, 58], with their consideration improving the accuracy of genetic risk predictions for several conditions [59–62].

Given that diseases rarely occur in isolation, understanding the etiology of diseases and their co-occurrence with others is critical for accurately assessing disease risk and developing effective prevention and treatment strategies at both global and personalized levels. These findings suggest that incorporating a broader range of factors—such as disease burden, environmental and lifestyle influences, and other omics data that capture dynamic molecular states closer to the phenotype (e.g., transcriptomics, epigenomics)—could significantly enhance the predictive accuracy of current risk models for individual and co-occurring diseases, ultimately leading to more tailored and effective interventions. An interesting conclusion is that most contributing factors to disease risk are a reflection of environmental exposures, and therefore, inherently malleable. This includes potentially heritable changes, such as certain epigenetic modifications [63], opening the door to new opportunities to disease and disease co-occurrence prevention and treatment.

The results suggest that gene expression is a critical layer in understanding the molecular underpinnings of disease co-occurrences, and its integration with genomic data provides near-complete coverage of DCs among the analyzed diseases, leaving less than 7% unexplained. This shows that almost all disease co-occurrences can be robustly captured with molecular information, with a high proportion likely rooted in factors beyond DNA sequence and, therefore, potentially modifiable. Furthermore, the data reveal that most comorbid diseases share robust systemic transcriptomic alterations, observable across different tissues (Note S4). This points to putative systemic alterations under a substantial portion of human diseases, a conclusion further supported by previous findings linking multimorbidity genes to broad tissue expression, high pleitropy [18] and systemic metabolic [13] or immune system functions [16].

Even though we observed that genomic recall is aligned with disease heritability, disease pairs may share additional genetic queues that were not detected in the UKB study due to insufficient sample size, population bias o the fact that the genetic cause depends on patient characteristics or the disease subtype. For this reason, larger studies including genomics and other kinds of patients’ molecular and clinical information will allow the community to disentangle the already proven molecular basis under most comorbidities. Longitudinal studies tracking gene expression and other molecular changes over time, alongside environmental exposures, could provide deeper insights into the dynamics of disease co-occurrence and progression. Furthermore, expanding the analysis to include more diverse populations will be essential to ensure that the findings are generalizable across different demographic groups, including variations in sex, age, or ethnicity.

## Materials and Methods

### Data collection and RNA-seq analysis

Publicly available and uniformly pre-processed RNA-seq data sets tackling human diseases were downloaded from the GREIN platform in June 2022 [64]. We removed the samples with less than 70% aligned reads to the genome and kept the studies that contained at least 3 cases and controls. This resulted in a dataset encompassing 148 diseases, derived from 107 studies, and comprising 6,512 samples. After aggregating the studies by disease, we followed the methodology described in Urda-Garćıa *et al*. to process the gene counts [16]. For each disease, we performed quality control, filtering of the lowly expressed genes, normalization, batch effect evaluation, and differential expression (considering case vs. control while correcting for the study of origin). Genes with a 5% FDR were considered significantly differentially expressed (sDEGs). Moreover, we applied Gene Set Enrichment Analysis (GSEA)[65] to obtain the diseases’ significantly differentially altered gene sets and pathways (5% FDR) from Reactome[66], Kyoto Encyclopedia of Genes and Genomes[67], and Gene Ontology[68].

### Construction of the RNA-seq networks

We built a **Disease Similarity Network (DSN)** by connecting diseases based on the significant similarities or dissimilarities of the differential gene expression profiles. Specifically, for each disease pair, we computed the cosine similarity between the logFC values of the genes in the union of their sDEGs. We used randomizations to assign a p-value to each disease pair and kept the significant pairs after multiple testing correction (1% FDR). In the DSN, two diseases are positively linked if they exhibit significantly similar differential gene expression profiles, and negatively linked if their profiles are significantly dissimilar.

Next, we followed the procedure described in Urda-Garćıa *et al*. to derive the **Stratified Similarity Network (SSN)**[16]. First, we stratified each disease into subgroups of patients with a similar gene expression profile (named meta-patients). Meta-patients were obtained by applying the k-medoids clustering algorithms to the normalized and batch effect corrected gene counts. Next, we characterized meta-patients in terms of their sDEGs. Finally, we built the SSN by connecting all phenotypes (diseases and meta-patients) based on the significant cosine similarity of their gene expression profiles (as previously described for the DSN). The SSN aims to encode the disease co-occurrences that are specific to some group of patients of a given disease.

We analyzed the topological properties of both the DSN and SSN, taking into account the entire networks and the subnetworks containing exclusively positive and negative interactions. When applicable, the topological properties were computed with and without considering the weight of the networks. For the negative subnetworks, the absolute value of the weight was considered. Moreover, properties requiring a distance as the edge weight were computed using 1 − cosine similarity + *ɛ*, where *ɛ* = 1 × 10*^−^*^8^.

### Comparison of the RNA-seq networks and UK Biobank interactions with the epidemiology

Firstly, we computed the overlap of the RNA-seq networks (DSN and SSN) with the gold-standard epidemiological network from Hidalgo *et al.*[5], containing comorbidities. To do so, we transformed our disease names into the International Code of Diseases version 9 (ICD9). For each RNA-seq network, we computed the overlap with the epidemiology (total number of captured comorbidities between the analyzed diseases), the recall (percentage of captured comorbidities between the analyzed diseases), and the precision (percentage of interactions in the network that are comorbidities). We assessed the significance of the recalls and precisions through randomizations (i.e. shuffling the interactions while preserving the degree distribution) (Table S3). Since the epidemiological network contains direct comorbidities, we computed the overlaps for the positive DSN and SSN subnetworks, containing positive interactions. Additionally, we evaluated the robustness of the recall to the removal of individual diseases and assessed the effect of imposing a threshold for the edge’s weight on the performance on the method. We also confirmed that the stratified network consistently improves the diseases’ recall without significantly affecting their precision.

Secondly, we followed the same procedure to evaluate the overlap of the DSN and SSN with the indenpendently generated epidemiological network from the UK Biobank (UKB) by Dong *et al.*, containing multimorbidities (Table S2) [18]. This time, disease names had to be transformed to the International Code of Diseases version 10 (ICD10) level 2. Since the UKB molecular layers only indicate the DCs in Dong *et al.* that could be explained by each data type, we can only compute their overlap and recall with Dong *et al*. (and not their precision or their comparison with Hidalgo *et al*). Analogously, the comparison of the different molecular layers has to be based on their overlap and recall from Dong’s *et al.* epidemiological network among the common set of diseases.

### Comparison of the molecular layers

We compared the ability of the different molecular layers (RNA-seq, SNPs, genes, pathways, PPIs, and genetic correlation) to overlap the disease co-occurrences in Dong *et al.* between their common diseases. We evaluated how these multimorbidities distribute between the different layers and their combinations. All computations were performed before and after filtering out the diseases that do not have GWAS summary statistics in the UKB.

To evaluate the dependency of the different layers, we used mutual information (the amount of information about one variable that you can get by observing the other one) [69]. Let I(X;Y) be the mutual information between variables X and Y and H(X), the entropy of X. We used I(X;Y)/H(X); i.e. the proportion of entropy of X explained by Y (Fig. 3c) to assess the directional dependency between the layers. In particular, we computed this metric for each pair of molecular layers, where each layer is characterized by a binary and equally sorted vector indicating if it captures Dong’s CDs. We also computed the normalized mutual information, Jaccard index, overlap (number of overlapping interactions), and recall between the different layers.

To quantify the relative contributions of genomic and transcriptomic layers, we introduced the Relative Recall Contribution (RRC) metric, which scales these values to a [−1, 1] linear range. The RRC is calculated as RRC = *<u>^G−T^</u>*, where *G* represents the recall of the genomic layers, and *T* represents the recall of the transcriptomics layers. This metric allows straightforward interpretation: when *T* = 100 and *G* = 0, the RRC is −1, indicating exclusive transcriptomics recall; when *T* = *G*, the RRC is 0, indicating balanced recall; and when *G* = 100 and *T* = 0, the RRC is +1, indicating exclusive genomic recall. Thus, the RRC offers a continuous scale for assessing relative recall contributions from genomic and transcriptomic layers. We applied quadratic regression to examine if diseases with a higher number of comorbidities exhibit RRC values near zero, indicating balanced contributions. To account for tied values, we used Kendall’s correlation to assess the association between the absolute RRC value and the number of disease co-occurrences. Additionally, Kendall’s correlation was used to assess the relationship between the number of co-occurrences and the individual recalls of the genomic layers.

### Heritability

We used recent ICD10 level 3 heritability (*h*^2^) estimates of human diseases derived from Danish nationwide registries spanning 40 years and covering a population of 6.3 million individuals [19]. To assess if diseases tend to co-occur with others of similar heritability, we applied assortativity—a metric capturing the tendency of nodes in a network to connect with those sharing specific attributes. Specifically, we computed assortativity by labeling diseases with their *h*^2^ values for the entire UKB epidemiological network and for the subnetworks containing epidemiological interactions supported by at least one molecular layer, at least one genomic layer, individual genomic layers, and gene expression. Additionally, we calculated the Spearman’s correlation between heritability and the proportion of DCs explained by each molecular layer to examine if disease heritability is associated with an increased likelihood of molecularly, genetically or transcriptomically-supported disease co-occurrences. We also examined the correlation between heritability and the ratio of DCs explained by gene expression versus genomics. This analysis was conducted for all diseases and for diseases with at least 3 DCs in the UKB epidemiological network.

## Data Availability

All data produced in the present work are contained in the manuscript.

## Acknowledgments

This work has been supported by the grants PRE2019-090454 funded by MICIU/AEI/10.13039/50110 0011033 and by “ESF Investing in your future”; and the grants RTI2018-096653-B-I00 and PID2022-141809OB-I00, both funded by MICIU/AEI/10.13039/501100011033 and by “ERDF/EU”, by the “European Union”. The research leading to these results has received funding from the European Community’s Horizon Europe Programme under grant agreement No. 101136957 (COMMUTE).

## Author Contributions

Beatriz Urda-Garćıa: Conceptualization, Formal Analysis, Investigation, Visualization, Writing-original draft, Writing-review & editing. Davide Cirillo: Conceptualization, Supervision, Writing-review. Alfonso Valencia: Conceptualization, Funding Acquisition, Supervision, Writing-review & editing.

## Data Availability

The code used for the analyses will be made available upon publication at https://github.com/beatrizurda/disentangling-disease-cooccurrences.git. The data is publicly available (Supplementary Data 1); the raw data can be downloaded from GEO (https://www.ncbi.nlm.nih.gov/geo/) and the counts can be downloaded from the GREIN platform (http://www.ilincs. org/apps/grein/).

