## Supplementary Information for "Disentangling the genetic and non-genetic origin of disease co-occurrences"

#### Notes

##### Note S1. Topological properties of the DSN and SSN

Both the DSN and the SSN consist of a single connected component, with a predominance of positive interactions (∼60%)(Table S1). As expected, the SSN also exhibits a higher degree, indicating a greater number of interactions per node compared to the DSN. On the other hand, the positive DSN and SSN display quite similar density, mean transitivity and unweighted mean distance, with approximately one-third of the potential edges present in both networks. This suggests a comparable level of overall connectivity between the two networks. Positive interactions exhibit higher transitivity than negative interactions in both networks, suggesting that positive interactions form more cohesive clusters.

##### Note S2. RNA-seq significantly recalls up to 81.7% of disease co-occurrences

We computed the overlap of the positive interactions in the Disease Similarity Network (DSN) and the Stratified Similarity Network (SSN) with the gold standard epidemiological network from Hidalgo *et al.*[1] and UK Biobank (UKB) epidemiological network by Dong *et al.*[2] (Methods).

The DSN significantly recalls 53.59% (p-value = 0) of the disease co-occurrences in Hidalgo *et al.* based on relative risks (RR) [1], reproducing the results shown in Urda-García *et al.*[3] with 330% more diseases (Table S2, S3). On the other hand, the SSN achieves an even higher and significant recall of 81.65% (p-value = 0.02), compared to the 64.1% of the previous SSN (127.4% increase). These recalls are robust to the removal of individual diseases even though diseases vary in their individual recalls. We confirmed that the stratified network consistently improves the diseases' recall without significantly affecting their precision (Fig. S2). We observed that imposing a threshold for the edge's weight does not enhance the performance of the method (measured as F1 score, i.e., the harmonic mean of recall and precision) for both the DSN and the SSN (Fig. S3, Note S5).

Next, we computed the overlap of the DSN and SSN with the independently generated epidemiological network from the UKB, achieving very similar and consistent results. The DSN significantly recalls 48.24% (p-value = 0.0098) and the SSN recalls 77.48% also significantly (p-value = 0.0153). This is additional evidence of the robustness of the recalls, maintained over considerably different epidemiological networks (Table S2). Specifically, Hidalgo *et al.* entails comorbidities for 3 million elderly patients from the USA[1] whereas the smaller UKB network contains multimorbidities among <400.000 UK individuals aged 40-69[2]. Therefore, we can conclude that the vast majority of disease co-occurrences (DCs) (∼80%) do have a molecular basis that can be consistently captured with disease subgroups' similarities based on gene expression profiles.

#### Note S3. UKB molecular layers

Dong *et al.* used the previously published UKB GWAS summary statistics [4] to infer the shared genomic components between the UKB multimorbidities (i.e., the UKB molecular layers) [2]. We briefly summarize and discuss how they were generated. All the details can be found in the original publications.

Firstly, Dong *et al.* removed the variants with minor allele frequency and selected the significant SNPs for each ICD10 code. Then, they used linkage disequilibrium to expand the disease-associated SNPs. They considered multimorbidity shared SNPs the ones associated to both diseases of the multimorbidity. Then, they inferred a series of molecular components in a hierarchical manner: genes, protein-protein interactions (PPIs), pathways, and genetic correlation.

Genes were considered disease-associated if they (1) directly containing a disease-associated SNP, (2) showed expression associated with a disease-associated SNP via eQTL, or (3) were identified through the MAGMA gene and gene-set aggregating tool. Multimorbidity shared genes are those associated with both diseases.

Multimorbidity shared PPIs were those in which one gene of the PPI is associated with one disease and the other gene is associated with the other disease. Next, they used the genes to obtain the disease-associated canonical pathways containing less than 200 genes from multiple collections (e.g. KEGG, Reactome). Multimorbidity shared pathways are those associated with one disease and containing at least one disease-associated gene of the other disease. Finally, they measured the genetic architecture similarity between the multimorbid disease pairs by computing their genetic correlation directly from the GWAS summary statistics. This informs about the dependence between the genetic influences of the diseases.

Overall, an inclusive approach was used to infer the genomic components and define when a multimorbidity shares them. On one hand, SNPs were expanded using linkage disequilibrium and disease-associated genes were expanded via the inclusion of eQTLs and gene aggregation techniques. Disease-associated pathways were amplified by incorporating multiple collections. Interestingly, multimorbid pathways use a permissive criterion in which pathways do not have to be associated with both multimorbid diseases to be considered a multimorbidity shared pathway. Instead, they have to be associated with one disease and at least one of its genes has to be associated with the other disease.

This generally expansive approach is expected to increase the amount of multimorbidities with shared genomic components. However, since Dong *et al.* do not provide the shared genomic components for the pairs of non-multimorbid diseases, we can not assess if multimorbidities share more genomic components than the expected by chance. Similarly, we can not assign a significance to the genomic layers' recall from the epidemiology nor can we compute their precision from it. This means that even if pathways underlie ∼30% of the UKB multimorbidities (being pathways the most informative genomic layer), this could be non-significant with respect to non-multimorbid diseases, which is possible given the permissive criterion that was used to define multimorbidity shared pathways. Conversely, we have shown that the transcriptomic layers do recall a significant proportion of the epidemiology and that they far exceed the ability of the genomic layers to recall multimorbidities, even under the explained context.

#### Note S4. Tissue of origin

In this work, we have exploited publicly RNA-seq studies tackling human diseases, where samples belong to individual diseases in a given tissue. Since each study includes both patient and control samples, we were able to correct for the tissue effect when generating sDEGs at the disease and stratified level. We observed diseases' systemic similarities that significantly and very robustly recalled the epidemiology regardless of the epidemiological network of reference, number, and specific subset of considered diseases. Thus, there is enough evidence to support that, even when different tissues are being compared, enough systemic effects prevail. This points to putative systemic alterations under a substantial portion of human diseases. This conclusion is further supported by the observation that multimorbidity genes tend to be expressed across a broader range of tissues and frequently exhibit high pleitropy [2]. Additionally, a considerable fraction of comorbidity-associated metabolic reactions are active across all tissues [5]. Core molecular processes, such as immune system alterations, have also been found to underlie a substantial fraction of disease co-occurrences [3]. Future comprehensive data sets on human diseases across an array of interesting tissues will allow to further examine this.

#### Note S5. Evaluation of edge filtering

We evaluated the performance of the method when imposing a threshold for the edges' weight. (Fig. S3). Specifically, we computed the recall, precision, and the F1 score (the harmonic mean of precision and recall) for the Disease Similarity Network (DSN) and the Stratified Similarity Network (SSN) over an array of thresholds for the edges' weight (Fig. S3a). Edges were filtered based on percentiles (if their weight was below an array of percentiles, left panels) or based on specific values (if their weight was below an array of specific values, right panels). We also show the total number of interactions and the total number of captured disease co-occurrences (DCs) for the resulting networks (Fig. S3b).

We observed that F1 score (the combined measure of recall and precision) decreases as the threshold becomes more stringent for both networks with percentiles and specific values. This indicates that filtering edges based on their weight does not improve the performance of the method. Indeed, precision increases very slowly and always entails a bigger decrease in recall. In fact, precision, recall, and the number of total and disease co-occurrences are fairly robust to the removal of edges, presenting very small changes unless a considerable portion of the network is removed (25-75%).

Importantly, epidemiological networks lack many true disease co-occurrences. They may miss the ones affecting a different population or underrepresented age or group, specific sex or disease subtypes, simply co-occurring within a larger time window than the one considered or without enough sample size. Hence, many interactions in our transcriptomic networks match DCs missed by Dong [2] or Hidalgo *et al.*[1] yet confirmed by other epidemiological studies with sufficient sample size or the appropriate set-up to observe the co-occurrence. For instance, many true DCs between neoplasms captured by our transcriptomic layers are missed by Hidalgo *et al.* because they tend to co-occur at a younger age than the one inspected in this study. Examples include the comorbidity relationships between breast cancer and lung, colorectal, thyroid cancer or Kaposi's sarcoma [6]. Therefore, we highlight that precision indicates the proportion of DCs in the transcriptomic networks described in a given epidemiological network, which is expected to lack many of the known DCs. In other words, many of the false positives that negatively affect precision might indeed be true positives missed by the epidemiological network of reference.

### Figures

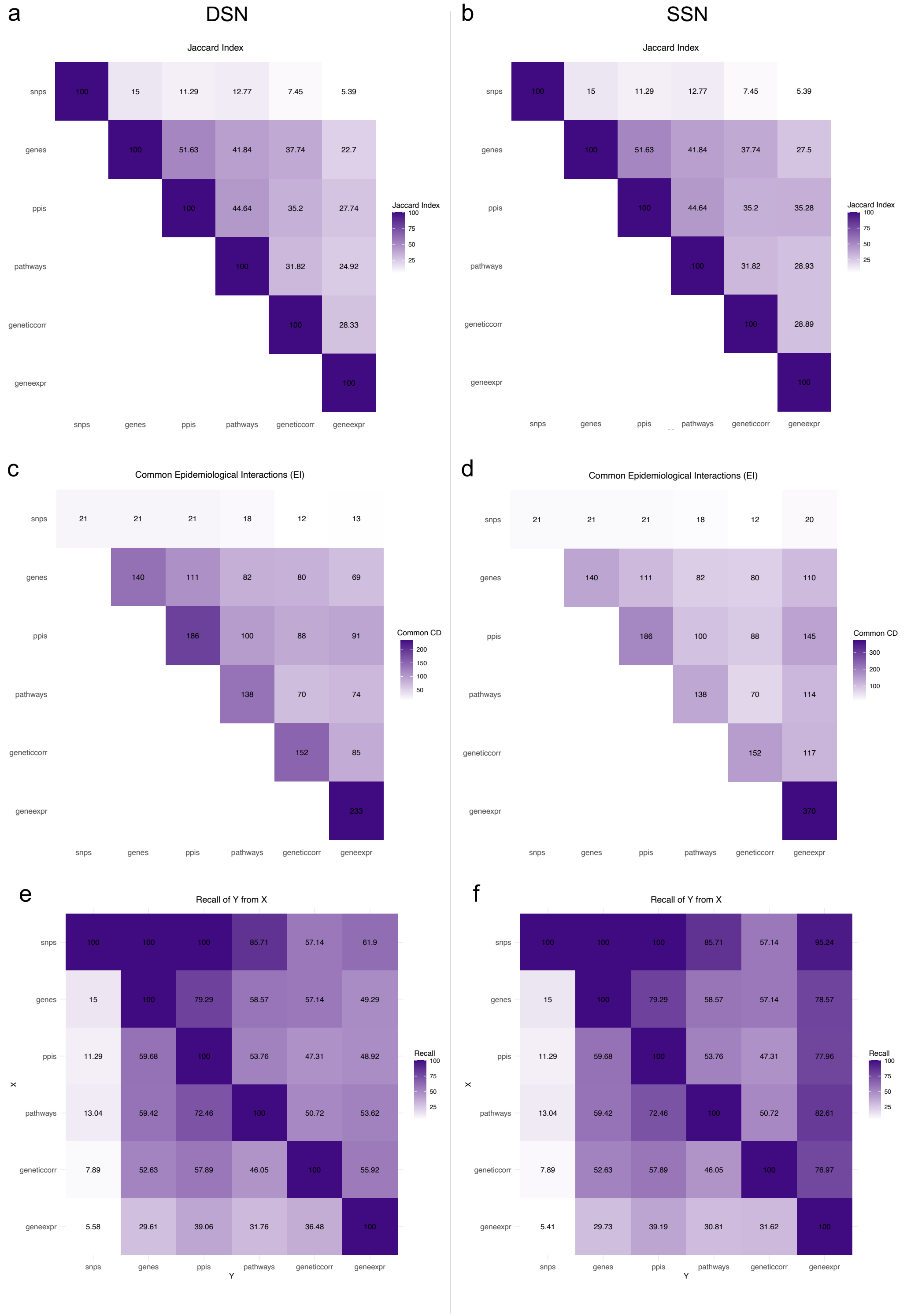


Figure S1: **Relationship between the different layers.** The gene expression layer corresponds to the Disease Similarity Network (DSN) in the left panel and to the Stratified Similarity Network (SSN) in the right panel. (a-b) Jaccard index between the disease co-occurrences (DCs) captured by each pair of layers over the common diseases containing GWAS. (c-d) Common number of DCs between each pair of layers. (e-f) Pairwise recall of all molecular layers. It shows the recall of variable Y (columns) of the disease co-occurrences explained by variable X (rows).

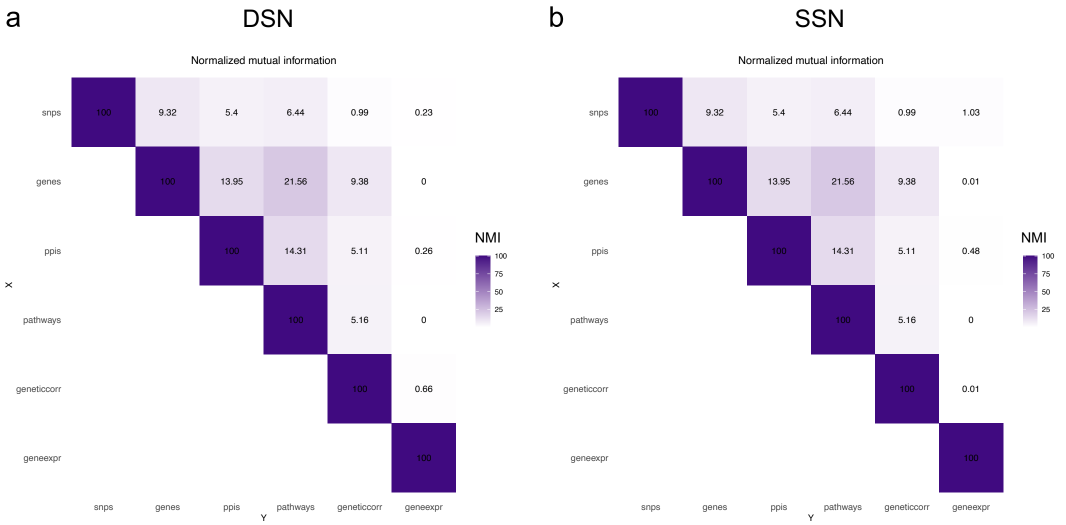


Figure S2: **Normalized mutual information between the molecular layers.** Heatmap containing the pairwise normalized mutual information between the molecular layers for (a) the Disease Similarity Network (DSN) and (b) the Stratified Similarity Network (SSN).

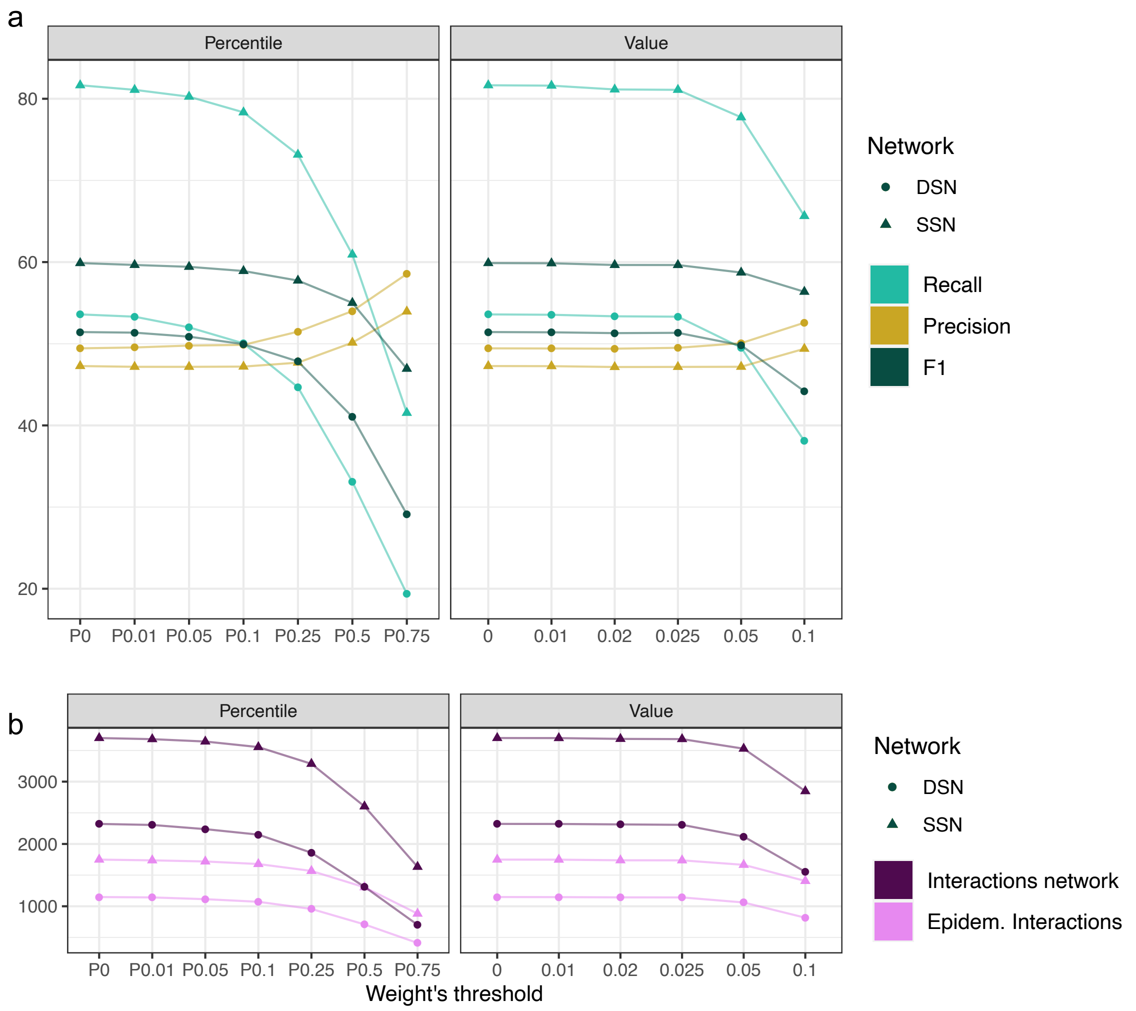


Figure S3: **Evaluation of edge filtering.** (a) It shows the recall, precision and F1 score of the Disease Similarity Network (DSN) and the Stratified Similarity Network (SSN) after imposing a threshold for the edge's weight based on the percentiles (Percentile) or specific values (Value) (Methods). (b) It shows the total number of interactions and the number of captured disease co-occurrences for each case.

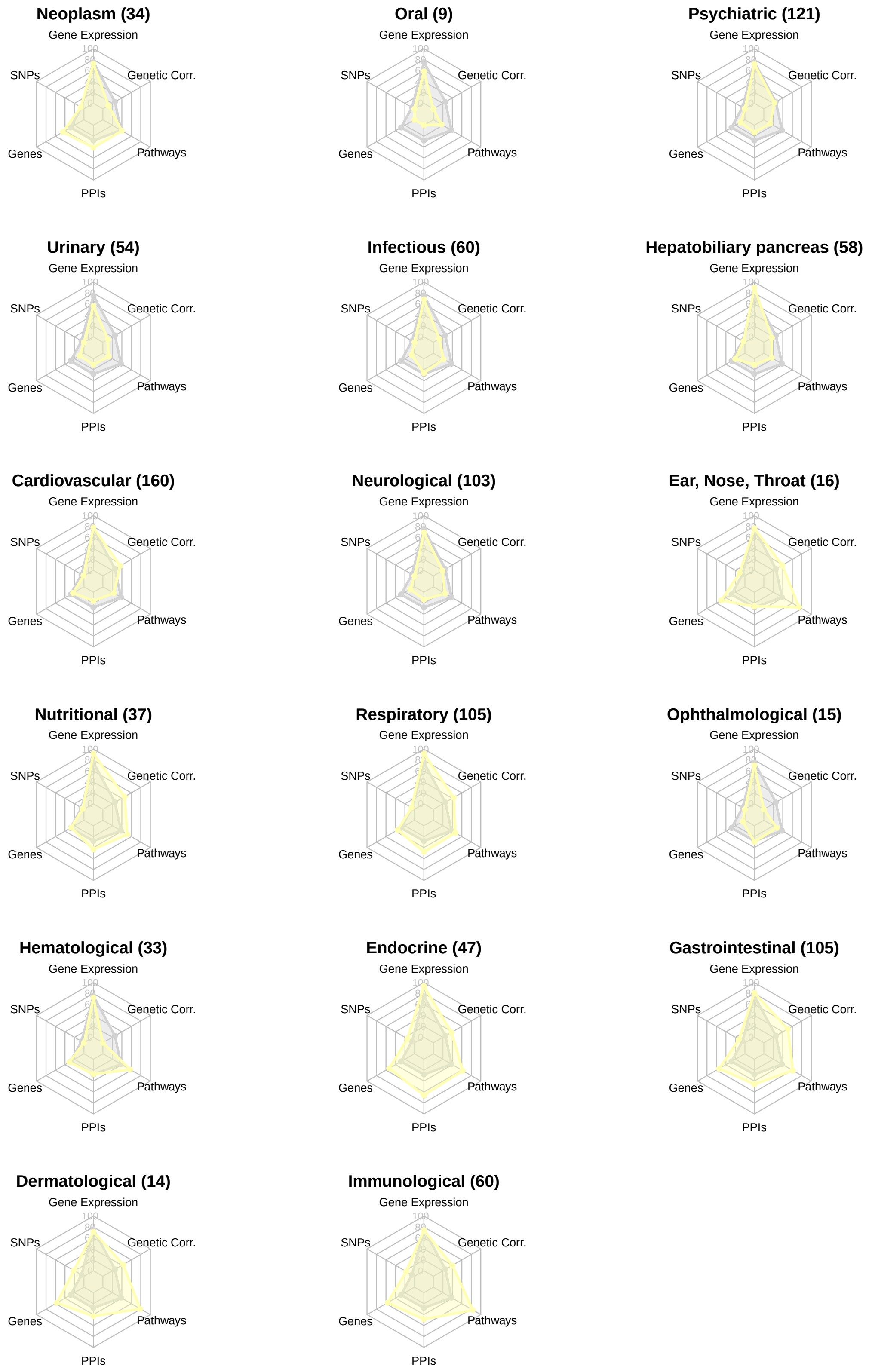
Neoplasm (34)
Gene Expression
Genetic Corr.
Pathways
PPIs
Genes
SNPs
Oral (9)
Gene Expression
Genetic Corr.
Pathways
PPIs
Genes
SNPs
Psychiatric (121)
Gene Expression
Genetic Corr.
Pathways
PPIs
Genes
SNPs
Urinary (54)
Gene Expression
Genetic Corr.
Pathways
PPIs
Genes
SNPs
Infectious (60)
Gene Expression
Genetic Corr.
Pathways
PPIs
Genes
SNPs
Hepatobiliary pancreas (58)
Gene Expression
Genetic Corr.
Pathways
PPIs
Genes
SNPs
Cardiovascular (160)
Gene Expression
Genetic Corr.
Pathways
PPIs
Genes
SNPs
Neurological (103)
Gene Expression
Genetic Corr.
Pathways
PPIs
Genes
SNPs
Ear, Nose, Throat (16)
Gene Expression
Genetic Corr.
Pathways
PPIs
Genes
SNPs
Nutritional (37)
Gene Expression
Genetic Corr.
Pathways
PPIs
Genes
SNPs
Respiratory (105)
Gene Expression
Genetic Corr.
Pathways
PPIs
Genes
SNPs
Ophthalmological (15)
Gene Expression
Genetic Corr.
Pathways
PPIs
Genes
SNPs
Hematological (33)
Gene Expression
Genetic Corr.
Pathways
PPIs
Genes
SNPs
Endocrine (47)
Gene Expression
Genetic Corr.
Pathways
PPIs
Genes
SNPs
Gastrointestinal (105)
Gene Expression
Genetic Corr.
Pathways
PPIs
Genes
SNPs
Dermatological (14)
Gene Expression
Genetic Corr.
Pathways
PPIs
Genes
SNPs
Immunological (60)
Gene Expression
Genetic Corr.
Pathways
PPIs
Genes
SNPs

Figure S4: **Radar charts of the molecular contributions to disease co-occurrences (DCs) of disease categories.** Each radar chart presents the ability of six distinct molecular layers —gene expression, genetic correlation, pathways, protein-protein interactions (PPIs), genes, and single nucleotide polymorphisms (SNPs)— in capturing the disease co-occurrences associated with a the diseases of a given disease category (yellow). The average values of all diseases are plotted in grey. The axes of the radar charts represent the recall of each molecular layer, indicating its ability to explain DCs. Longer axis lengths signify a higher recall, suggesting a greater influence of the corresponding molecular layer on the DCs of the disease category. ICD10 disease categories are sorted based on the ratio of DCs explained by transcriptomics versus genomics, from high to low. The number of interactions in UKB epidemiological network (NDCs) for each disease category is shown between parentheses.

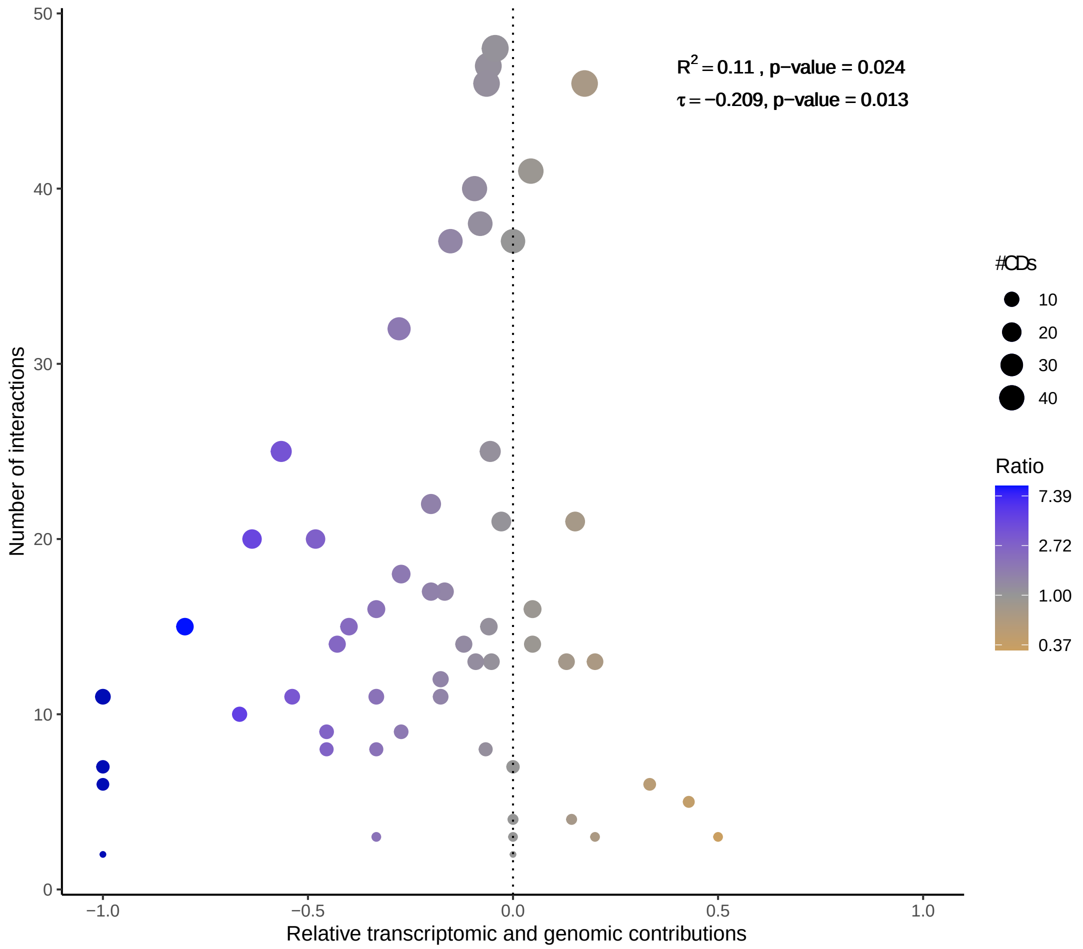


Figure S5: **Number of disease co-occurrences and relative molecular contributions.** Scatter plot showing the relationship between the number of disease co-occurrences (DCs) and the relative genomic and transcriptomic contributions to DCs of human diseases. Each point represents a disease, with the y-axis indicating the number of co-occurrences in the UKB epidemiological network within the disease set. The x-axis shows the Relative Recall Contribution (RRC), which quantifies the balance between genomic and transcriptomic contributions on a $[-1, 1]$ scale. An RRC of $-1$ denotes exclusive transcriptomic contributions, 0 indicates balanced contributions, and $+1$ denotes exclusive genomic contributions (Methods). Point size reflects the number of co-occurrences, and color indicates the ratio of transcriptomic to genomic recall, with yellow for diseases better explained by genomics and blue for those better explained by transcriptomics. Quadratic regression analysis shows that diseases with a higher number of comorbidities tend to have RRC values near zero, suggesting balanced molecular contributions ($\mathbf{R^2}$ = 0.11, p-value = 0.024). To account for tied values, Kendall's correlation ($\tau$ = -0.209, p-value = 0.013) was used to evaluate the association between the absolute RRC value and the number of disease co-occurrences (Methods).

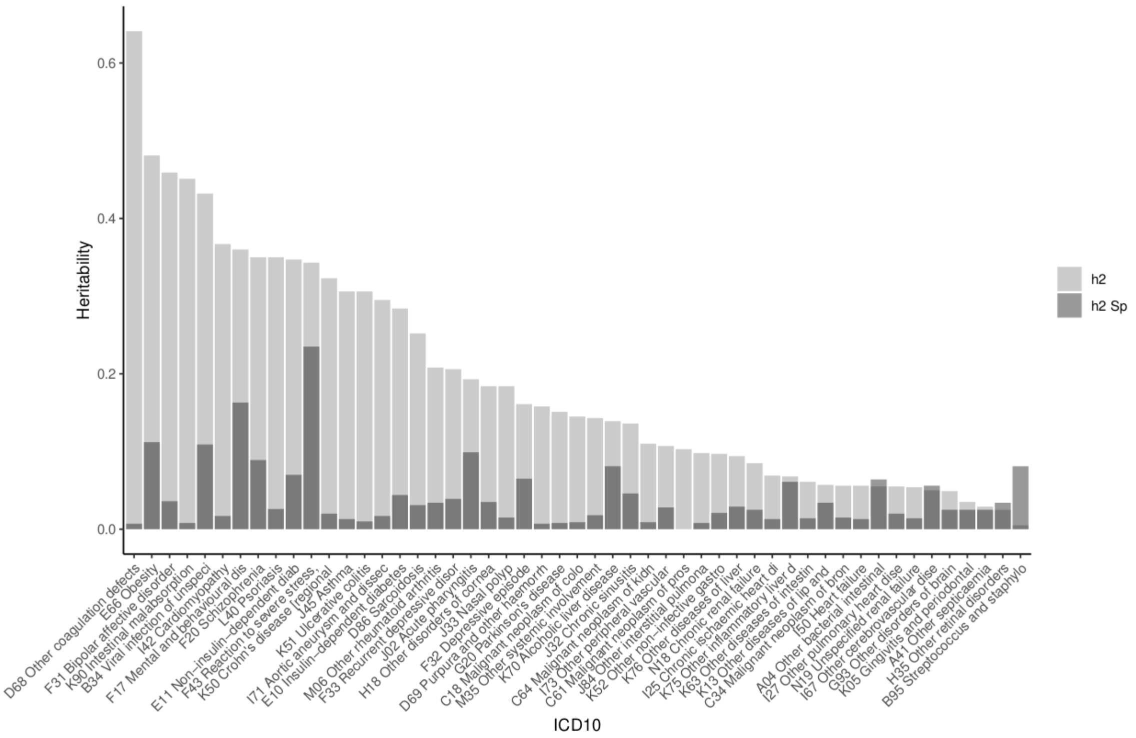


Figure S6: **Heritability and spousal heritability of human diseases.** Heritability (h2) and spousal heritability (h2 Sp) of the considered diseases with genomic information from the UKB measured by [7] (Methods). Light grey bars denote the heritability (h2) values, while dark grey bars represent the spousal heritability (h2 Sp) values. Diseases are sorted by their h2 values and labeled by their ICD10 code and disease name. The plot displays the genetic and environmental contributions shared with spouses to diseases.

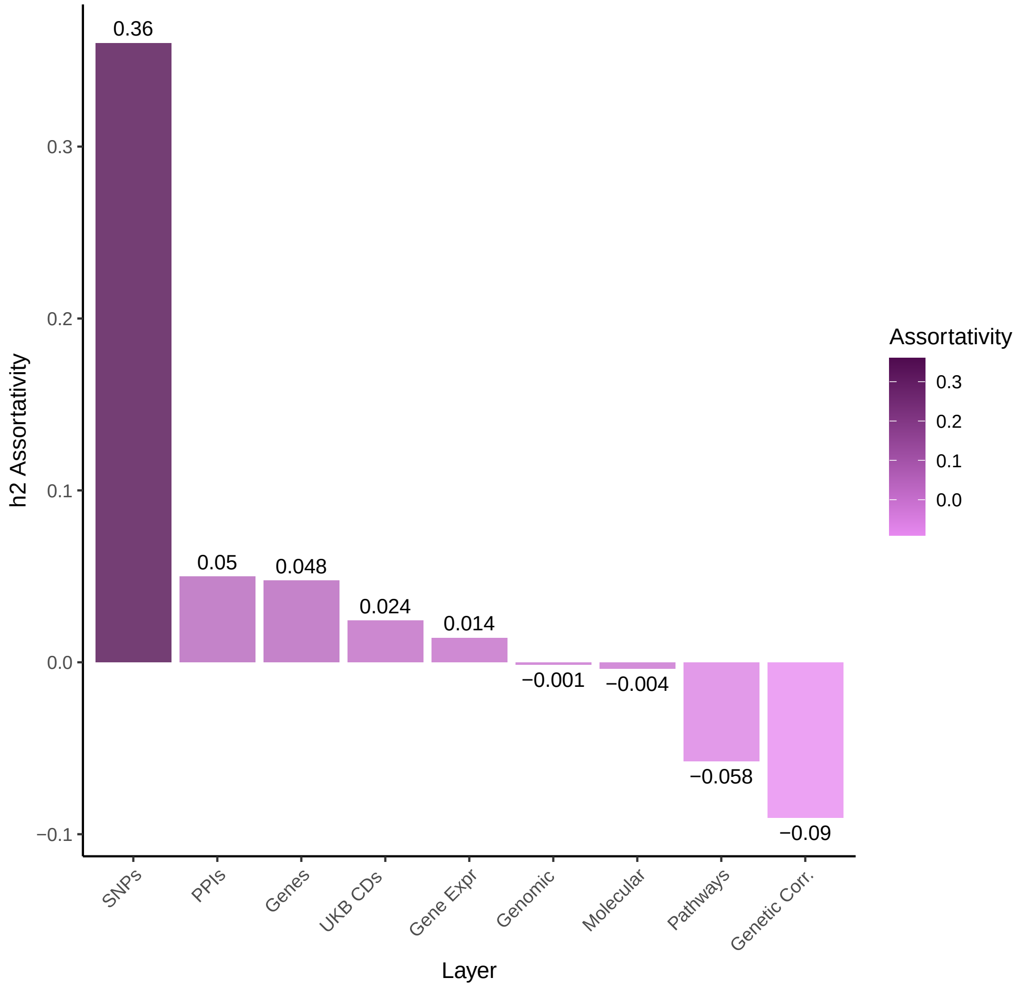


Figure S7: **Heritability assortativity of disease co-occurrences.** Bar plot showing the assortativity of different layers in terms of heritability (h2), ordered from highest to lowest. Assortativity is a measure of the tendency of nodes in a network to connect to other nodes that are similar in some way. Therefore, h2 Assortativity measures the tendency of diseases (nodes) to connect to diseases with similar heritabilities. Layers include all disease co-occurrences in the UKB epidemiological network (UKB DCs) and the subnetworks recalled by SNPs, Genes, PPIs, Pathways, Genetic Correlation, Gene Expression, Genomic (any genomic layer), and Molecular (any molecular layer). The SNPs subnetwork is the only one where diseases tend to link to diseases with similar heritabilities.

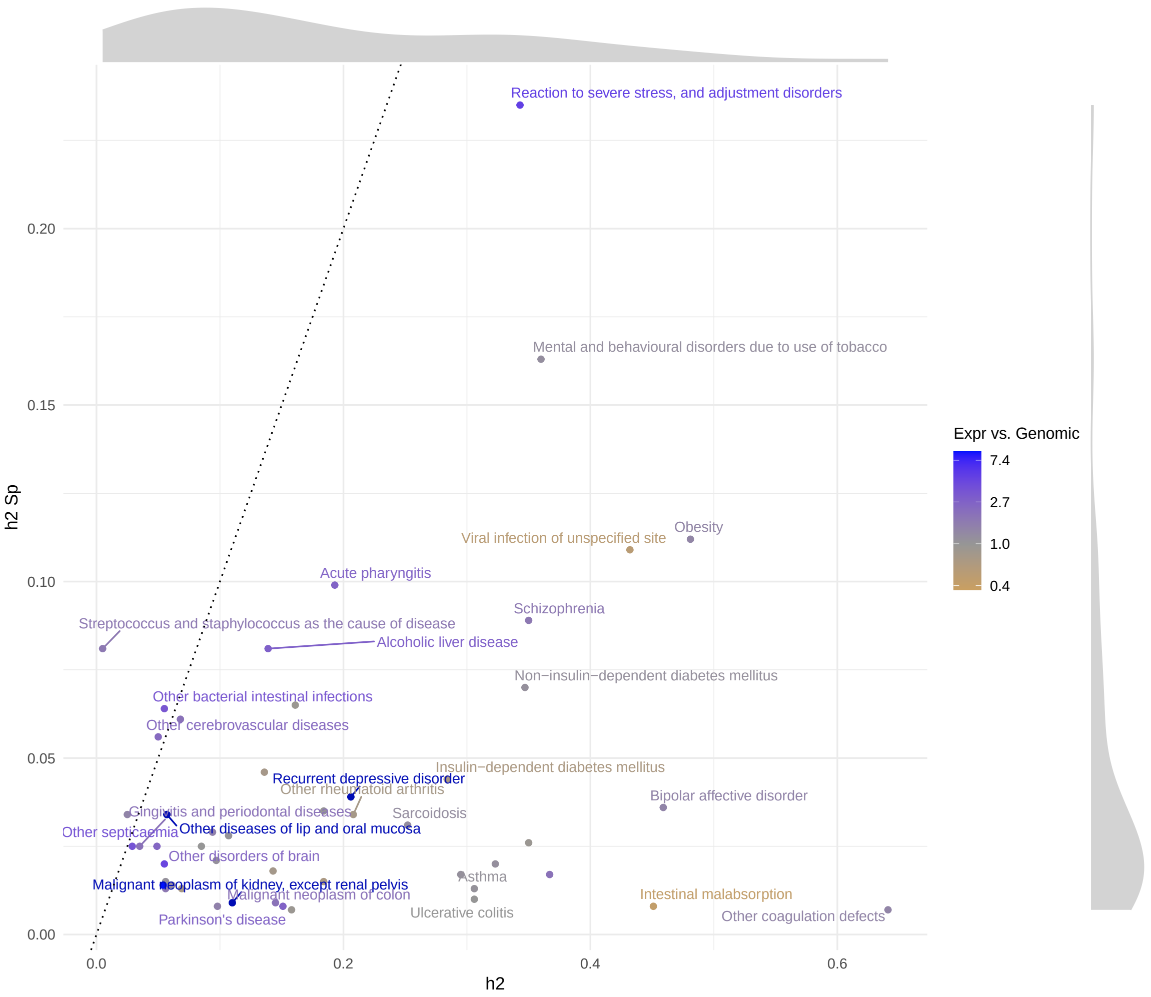


Figure S8: **Spousal heritability and heritability of human diseases.** Scatter plot displaying the Spousal heritability (h2 Sp) versus the heritability (h2) of human diseases. The diseases are colored based on the ratio between the proportion of disease co-occurrences (DCs) captured by transcriptomics versus genomics. Blue indicates diseases where co-occurrences are better captured by transcriptomics, while yellow indicates diseases where co-occurrences are better captured by genomics. The distributions of h2 and h2 Sp values are shown in grey at the top and right side of the figure, respectively. The dotted identity line represents points where h2 equals h2 Sp.

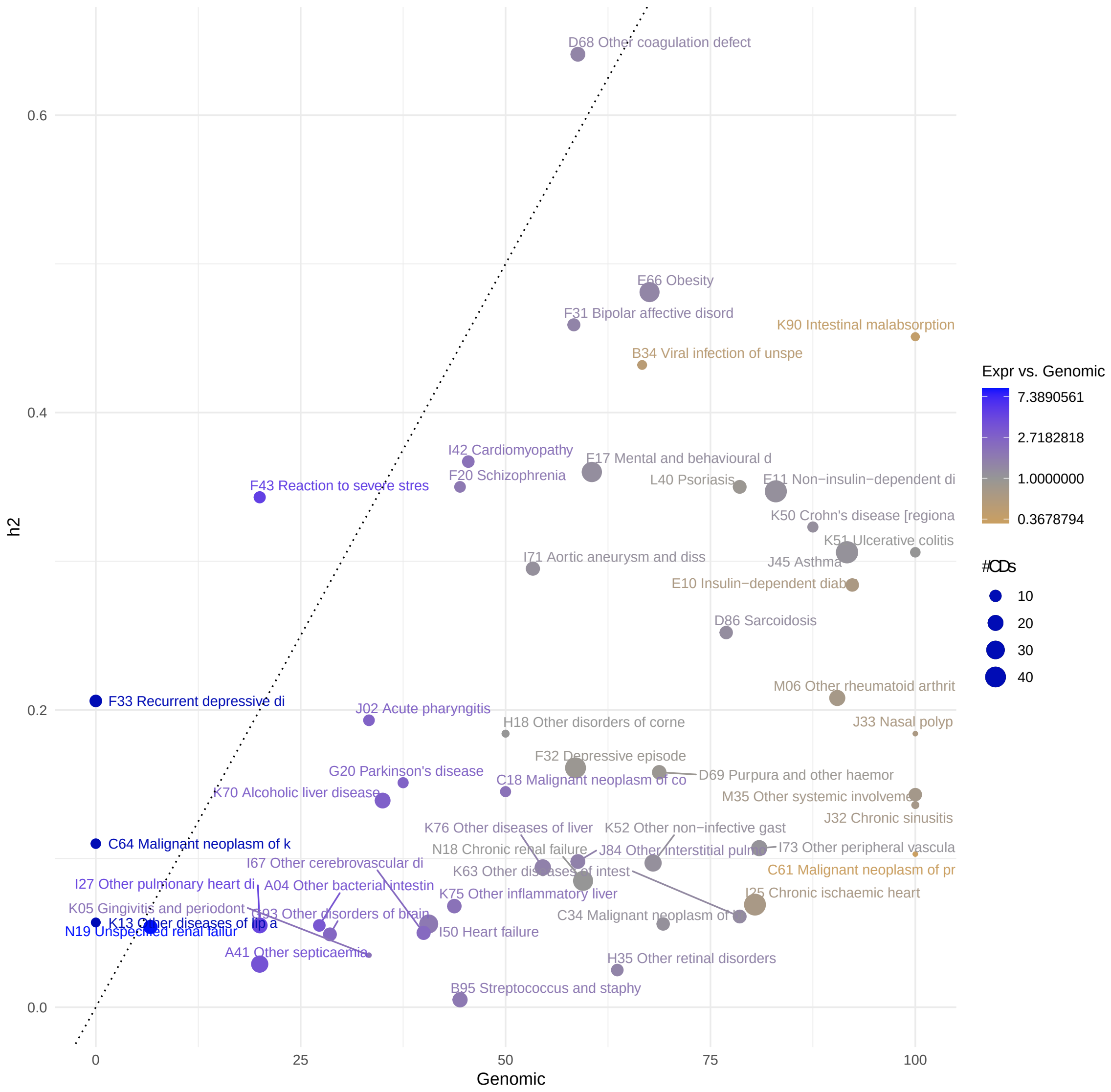


Figure S9: **Heritability of human diseases and DCs genomic recall.** Scatter plot displaying the heritability (h2) versus the proportion of disease co-occurrences (DCs) captured by genomics of human diseases. Point size corresponds to the number of DCs associated with each disease in the UK Biobank epidemiological network [2]. Diseases better explained by genomics are depicted in blue, while those better explained by transcriptomics are shown in yellow. Dark blue points represent diseases whose DCs are exclusively explained by transcriptomics.

### Tables

Table S1: **Topological properties of the transcriptomic networks.**
Table containing the topological properties of the Disease Similarity Network (DSN) and the Stratified Similarity Network (SSN) containing all or only positive or negative interactions. It shows the number of connected components, number of nodes, number and percentage of total, positive and negative interactions, as well as the topological properties of these networks. When applicable, the topological properties have been computed with and without considering the weight of the networks (unweighted). The weight of the edge corresponds to the cosine similaritiy between the differential gene expression profile of the diseases. For the negative subnetworks, the absolute value of the weight has been considered. Moreover, properties requiring a distance as the edges' weight use one minus the weight (Methods).

| | **DSN** | | | **SSN** | | |
|---|---|---|---|---|---|---|
| Interaction type | **All** | **Positive** | **Negative** | **All** | **Positive** | **Negative** |
| Connected components | 1 | 1 | 1 | 1 | 1 | 1 |
| Number of nodes | 148 | 148 | 148 | 511 | 511 | 511 |
| Number of interactions | 5954 | 3681 | 2273 | 72286 | 42148 | 30138 |
| Positive interactions | 3681 | 3681 | 0 | 42148 | 42148 | 0 |
| Negative interactions | 2273 | 0 | 2273 | 30138 | 0 | 30138 |
| % positive interactions | 61.82 | 100 | 0 | 58.31 | 100 | 0 |
| % negative interactions | 38.18 | 0 | 100 | 41.69 | 0 | 100 |
| Mean degree | 80.46 | 49.74 | 30.72 | 282.92 | 164.96 | 117.96 |
| Edge density | 0.55 | 0.34 | 0.21 | 0.55 | 0.32 | 0.23 |
| Transitivity | 0.61 | 0.51 | 0.16 | 0.63 | 0.5 | 0.16 |
| Diameter | 1.74 | 2.14 | 2.53 | 1.6 | 1.77 | 2.24 |
| Diameter (unweighted) | 2 | 3 | 3 | 2 | 3 | 3 |
| Mean closeness | 4.75e-03 | 4.12e-03 | 3.77e-03 | 1.38e-03 | 1.18e-03 | 1.11e-03 |
| Mean betweenness | 71.74 | 90.77 | 65.18 | 853.39 | 866.48 | 338.18 |
| Mean betweenness (unweighted) | 33.27 | 48.81 | 59.7 | 113.54 | 172.56 | 197.04 |
| Mean distance | 0.94 | 1.08 | 1.39 | 0.73 | 0.83 | 1.17 |
| Mean distance (unweighted) | 1.45 | 1.66 | 1.81 | 1.45 | 1.68 | 1.77 |

Table S2: **Overlap between the RNA-seq networks and the epidemiology.** Table showing the overlap of the Disease Similarity Network (DSN) and Stratified Similarity Network (SSN) constructed from RNA-seq data with epidemiological networks by Hidalgo *et al.*[1] and Dong *et al.*[2]. For the Hidalgo *et al.* networks, results are shown separately for networks based on relative risks (RR), phi-correlation, and the combination of both approaches. For each case, the number of overlapping disease co-occurrences (Overlap), Recall, and Precision are reported along with their corresponding p-values (columns) (see Methods).

| **Reference** | **Network** | **Overlap** | **Recall (%)** | **p-value** | **Precision (%)** | **p-value** |
|---|---|---|---|---|---|---|
| Hidalgo *et al.* RR | DSN | 1148 | 53.59 | 0 | 49.44 | 0 |
| | SSN | 1749 | 81.65 | 0.02 | 47.27 | 0.0166 |
| Hidalgo *et al.* phi-correlation | DSN | 1584 | 51.55 | 0.0001 | 68.22 | 0 |
| | SSN | 2475 | 80.54 | 0.0077 | 66.89 | 0.004 |
| Hidalgo *et al.* combined | DSN | 1584 | 51.55 | 0.0001 | 68.22 | 0 |
| | SSN | 2475 | 80.54 | 0.0077 | 66.89 | 0.004 |
| Dong *et al.* | DSN | 302 | 48.24 | 0.0098 | 24.06 | 0.032 |
| | SSN | 485 | 77.48 | 0.0153 | 24.00 | 0.0474 |

Table S3: **Overlap between the RNA-seq networks and the epidemiological network by Hidalgo *et al.***
Table containing the overlap of the Disease Similarity Network (DSN) and Stratified Similarity Network (SSN) from the epidemiological network by Hidalgo *et al.*, based on relative risks in the previous study by Urda-García *et al.* (Original) and in this new study (New). For each case, the number of overlapping disease co-occurrences (Overlap), Recall, and Precision are reported along with their corresponding p-values (Methods).

| Data set | Network | Overlap | Recall (%) | p-value | Precision (%) | p-value |
|---|---|---|---|---|---|---|
| Original | DSN | 162 | 49.24 | 0.0034 | 44.38 | 0.0021 |
| | SSN | 211 | 64.13 | 0.0187 | 43.06 | 0.0253 |
| New | DSN | 1148 | 53.59 | 0 | 49.44 | 0 |
| | SSN | 1749 | 81.65 | 0.02 | 47.27 | 0.0166 |

Table S4: **Correlation between the proportion of DCs captured by molecular layers and heritability.** For each disease, we computed the proportion of disease co-occurrences (DCs) captured by the molecular layers: SNPs, genes, PPIs, pathways, genetic correlation, any genomic layer (genomic), and gene expression. Additionally, we calculated the ratio of DCs captured by gene expression relative to genomic layers (Gene expression vs. genomic). This table shows the Spearman's correlation and corresponding p-value for the association between the proportion of DCs captured by each molecular layer (rows) and the heritability of the diseases (Methods). The analysis is conducted for diseases with at least 3 DCs and for all diseases (columns). The results indicate that the proportion of DCs captured by most genomic layers significantly correlates with heritability, suggesting that diseases with higher heritability tend to have DCs that are better captured by genomic information compared to less heritable diseases. This trend is not observed for gene expression.

| | **Diseases >3 DCs** | | **All diseases** | |
|---|---|---|---|---|
| **Layer** | **Correlation** | **p-value** | **Correlation** | **p-value** |
| SNPs | 0.34* | 0.016 | 0.31* | 0.026 |
| Genes | 0.25 | 0.079 | 0.19 | 0.17 |
| PPIs | 0.48* | 0.00033 | 0.42* | 0.0019 |
| Pathways | 0.28* | 0.046 | 0.27* | 0.047 |
| Genetic correlation | 0.38* | 0.0058 | 0.33* | 0.015 |
| Genomic | 0.37* | 0.0072 | 0.34* | 0.013 |
| Gene expression | 0.048 | 0.74 | -0.0077 | 0.96 |
| Gene expression vs. genomic | -0.36* | 0.011 | -0.37* | 0.0086 |

Table S5: **Top 15 diseases based on spousal heritability.**
Table containing the top 15 diseases included in this study with the highest Spousal heritability (h2 Sp) in [7]*et al.* It also shows the proportion of disease co-occurrences captured by gene expression (Gene Expr), the proportion of DCs captured by gene expression versus genomics (Ratio), and the ICD10 disease category.

| **Disease name** | **h2 Sp** | **Gene Expr.** | **Ratio** | **Disease category** |
|---|---|---|---|---|
| F43 Reaction to severe stress, and adjust.. | 0.235 | 100.00 | 5.00 | Psychiatric |
| F17 Mental.. disorders due to.. tobacco | 0.163 | 71.05 | 1.17 | Psychiatric |
| E66 Obesity | 0.112 | 91.89 | 1.36 | Nutritional |
| B34 Viral infection of unspecified site | 0.109 | 33.33 | 0.50 | Infectious |
| J02 Acute pharyngitis | 0.099 | 88.89 | 2.67 | Ear, Nose, Throat |
| F20 Schizophrenia | 0.089 | 77.78 | 1.75 | Psychiatric |
| B95 Streptococcus and staphylo. | 0.081 | 77.78 | 1.75 | Infectious |
| K70 Alcoholic liver disease | 0.081 | 100.00 | 2.86 | Hepatobiliary pancreas |
| N02 Recurrent and persistent haematuria | 0.079 | 50.00 | Inf | Urinary |
| E11 Non-insulin-dependent diabetes... | 0.07 | 93.62 | 1.13 | Endocrine |
| F32 Depressive episode | 0.065 | 53.66 | 0.92 | Psychiatric |
| A04 Other bacterial intestinal infections | 0.064 | 90.91 | 3.33 | Infectious |
| K75 Other inflammatory liver diseases | 0.061 | 87.50 | 2.00 | Hepatobiliary pancreas |
| I67 Other cerebrovascular diseases | 0.056 | 93.33 | 2.33 | Cardiovascular |
